# Larger social networks may increase stigma against vocal illness: An integrated empirical and computational study of deciphering help-seeking behaviors and vocal stigma

**DOI:** 10.1101/2023.12.08.23299730

**Authors:** Aaron R. Glick, Colin Jones, Lisa Martignetti, Lisa Blanchette, Theresa Tova, Allen Henderson, Marc D. Pell, Nicole Y. K. Li-Jessen

**Affiliations:** School of Communication Sciences and Disorders, Faculty of Medicine and Health Sciences, McGill University, Quebec, Canada; School of Medicine, University of Montreal, Quebec, Canada; The Alliance of Canadian Cinema, Television and Radio Artists, Toronto, Ontario, Canada; National Association of Teachers of Singing, Jacksonville, Florida, U.S.A.; The Centre for Research on Brain, Language and Music, McGill University, Montreal, Quebec, Canada; Department of Biomedical Engineering, McGill University, Montreal, Quebec, Canada; Department of Otolaryngology - Head and Neck Surgery, McGill University, Montreal, Quebec, Canada; Research Institute of McGill University Health Center, Montreal, Quebec, Canada; Institute of Child Development, University of Minnesota, Minneapolis, Minnesota, USA; Masonic Institute for the Developing Brain, University of Minnesota, Minneapolis, Minnesota, USA

**Keywords:** agent-based models, health stigma, social computing, social networks, vocal health

## Abstract

**Background:** Individuals with a stigmatized medical condition are often reluctant to seek medical help. Among professionals, singers and actors often experience stigma associated with voice disorders. Scientific evidence for vocal stigma is, however, limited and primarily anecdotal. No quantitative research has explored the impact that vocal stigma may have on help-seeking behavior in professional vocal performers. This study deployed and integrated empirical and computational tools to (1) quantify the experience of vocal stigma and help-seeking behaviors and (2) predict their modulations with peer influences in social networks.

**Methods:** Experience of vocal stigma and information-motivation-behavioral (IMB) skills were prospectively profiled using online surveys from a total of 403 Canadians (200 vocal performers and 203 controls). The survey data were used to formulate an agent-based network model that numerically simulates the effect of social interactions on vocal stigma and help-seeking behaviors. Each virtual agent updates their IMB states via social interaction, which in turn changes their self- and social-stigma states. Profiles from vocal performers and non-vocal performers were compared as a function of network size. Network analysis was performed to evaluate the effect of social network structure on the flow of information and motivation among virtual agents.

**Results:** Over 4000 simulation runs in each context, larger social networks are more likely to contribute to an increase in vocal stigma. For small social networks, total stigma was reduced with higher total IMB but much less so for large networks with around 400 agents. For the agent population of vocal performers with high social-stigma and risk for voice disorder, their vocal stigma is resistant to large changes in IMB. Agents with extreme IMB and stigma values are also likely to polarize their networks faster in larger social groups.

**Conclusions:** We used empirical surveys to contextualize vocal stigma and IMB in real world populations and developed a computational model to theorize and quantify the interaction among stigma, health-seeking behavior and influence of social interactions. This work establishes an effective, predictable experimental platform to provide scientific evidence in developing public policy or social interventions of reducing health stigma in voice disorders and other medical conditions.

## Background

Occupational voice users contribute a significant labour force in Canada and elsewhere [1–5]. For instance, more than 35,000 professional artists such as voice performers, radio hosts and singers are employed in Canada as of 2016 [6]. Alarmingly, the estimated lifetime prevalence of voice problems is as much as 80% for occupational voice users [7–9]. They also report devastating physical and emotional difficulties as well as challenges in professional and career development [7, 8, 10, 11]. The direct health care cost associated with voice problems is estimated as $1.75 billion/year in Canada [12, 13]. Indirect health care costs related to anxiety, loss of workdays and employment, and other negative quality-of-life sequelae are incalculable [14–18]. Reduction in occupational vocal problems would improve patient well-being and reduce the healthcare burden on society.

### Theoretical Framework of Health Stigma

Stigma is considered a serious threat to population health inequalities as it disrupts resource availability, social relationships and coping behaviors within society [19]. Individuals with a stigmatized medical condition may become reluctant to seek medical help [20]. For example, a meta-analysis on mental health stigma found a negative correlation between stigma and help-seeking [21]. Similarly, a meta-analysis of stigma and health outcomes in HIV/AIDS found that those who experience symptoms of stigma were 21% less likely to access health and social services [22]. Individual studies also found associations between stigma and avoidance or delaying of help-seeking in other health issues, including alcoholism [23] and cancers [24]. Similarly, stigma against vocal illness, i.e., *vocal stigma*, is a known barrier to vocal performers seeking help for work-related illness.

In 2019, Stangl et al. published a seminal paper “*The Health Stigma and Discrimination Framework*” in BMC Medicine [25]. The paper proposed a hierarchy of social levels on which stigma operates, and a series of processes leading from the creation of a stigma to its measurable outcomes. In brief, the proposed framework consists of a multi-level social hierarchy, namely, individual, interpersonal, organizational, community, and public policy. Within this hierarchy, the stigmatization process starts with constituent factors that drive or facilitate health stigma. Then, individuals with a specific health condition are “marked” or “labeled” with a stigma, followed by manifestations, i.e., attitudes, behaviors, and experiences that arise from the stigma in those who are marked by the stigma and those around them. Finally, the stigma manifestations affect a range of measurable outcomes on affected populations (e.g., access to healthcare) and the society (e.g., public policies against discrimination).

By adopting Stangl et al.’s framework to vocal stigma, vocal performers with vocal impairments can entail catastrophic fears about losing the employment, social status, and creative fulfilment that their art provides [26]. In other words, they fear the consequences of not being able to meet the vocal demands of performance. This fear is a strong candidate for a driving factor in vocal stigma among vocal performers. However, individuals face different vocal demands and consequences for being unable to perform. For example, actors and singers face different vocal demands, and full-time performers face greater consequences from voice disorders than those who have other sources of income. (**Figure 1**)

**Figure 1.**
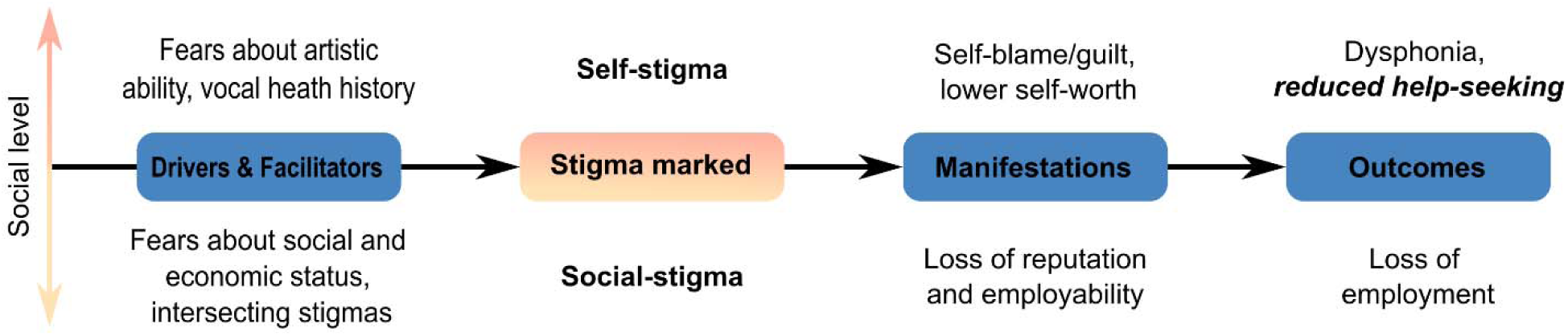
Proposed Health Stigma and Discrimination Framework for Vocal Stigma. Adapted from Stangl et al. (2019)

### Predicting Help-Seeking Behavior

Predicting help-seeking and other behaviors that influence health is a major topic in the field of health psychology [27]. One prominent psychological model for making these predictions is the Information Motivation Behavioral Skills (IMB) model [28]. *Information* represents a person’s knowledge and beliefs about the behavior, and about the issue that the behavior is intended to address. *Motivation* represents a person’s attitudes toward the predicted behavior, and their perception of social norms around it, i.e., what attitudes does the person believe other people hold? An example can be perceived costs and benefits of the behavior. *Behavioral Skills* represents a person’s ability to perform the behavior. This component can include objective skills, psychological skills and more importantly self-efficacy, i.e., a person’s intention to seek help [29].

By applying this IMB model to vocal stigma, *information* includes a person’s knowledge and belief about voice disorders and related issues. *Motivation* includes a person’s attitudes and perception of social norms about seeking professional medical help for a voice disorder. Finally, *behavioral skills* represent the intention to seek professional medical help if they acquire a voice disorder (**Figure 2**).

**Figure 2.**
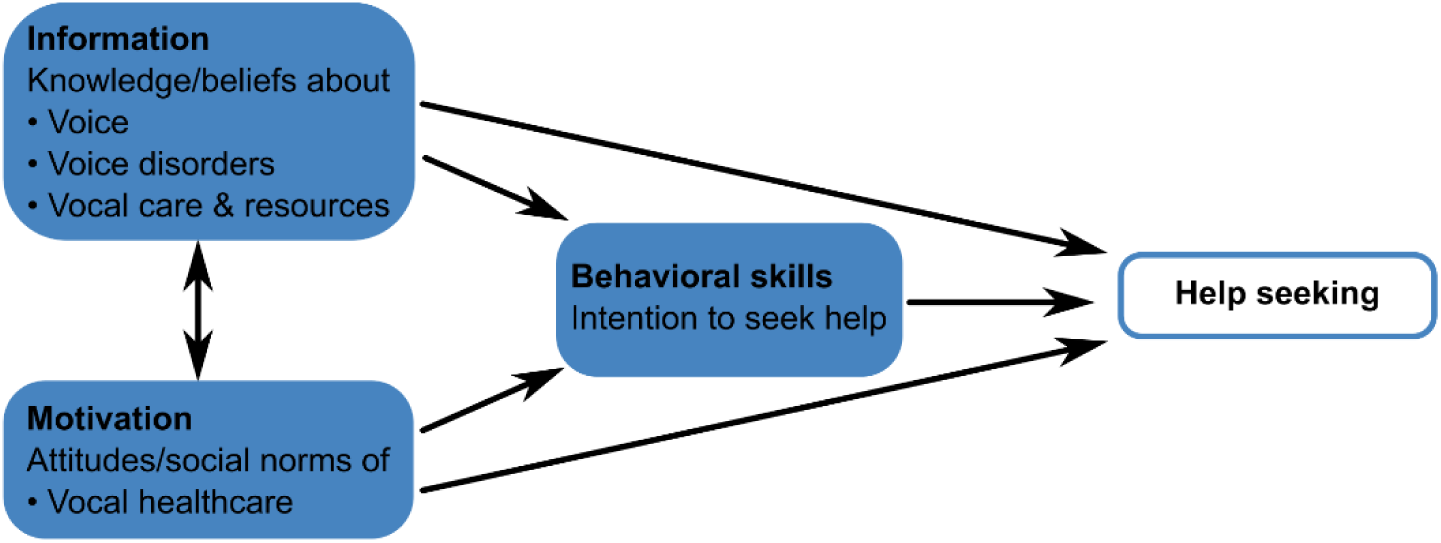
Proposed Information Motivation Behavioral Skills Model of Predicting Help-Seeking Behavior for Voice Disorders. Adopted from Fisher et al. (2002).

### Computer Simulation of Social Networks

In response to the well-recognized health challenges presented by stigma, various strategies have been proposed to reduce the stigma around conditions such as HIV and mental illness. At the level of social-stigma, one common approach is to provide information about a health condition to the general public, emphasizing that affected individuals are not to blame for their condition. Other interventions includ promoting empathy and understanding toward affected individuals, for example via testimonials or social contact between affected and unaffected individuals. For self-stigma, a provision of counselling or coping strategies can also be implemented to affected individuals [30–32].

Of these, social contact has emerged as the most successful strategy for reducing stigma in adults [30, 33]. Computer simulation has emerged as a powerful, quantitative way to study social networks and their effects on human behavior and public policy. In fact, social networks have been investigated as a structural determinant of health on their role in the spread of social behaviors and attitudes affecting health outcomes [34]. For instance, testable models have been developed to simulate the mechanisms of misinformation, bias, and polarization in social networks [35, 36], characterize the effects of misinformation during the COVID-19 pandemic [37], and help understand stigma and its impact on obesity, depression, mental illness, heart disease, and HIV-prevention [38–41].

In particular, agent-based models (ABM) have become a pivotal simulation platform in social and behavioral sciences in the fast-growing field of *social computing* [42–44]. ABM have been applied for building and testing theories in stock markets [45–47], consumer behaviors [48–53], and psychological cooperation [54–57]. For driver behaviors in particular [58–66], ABMs are used to simulate how the behavior of a driver (*an individual agent*) will affect other drivers (*other agents*) and the evolving traffic patterns (*emergent population behavior*). Rather than individual isolated decisions, social behaviors often resulted from interactions among people with diverse backgrounds over time. Such characteristics match very well with the framework of ABMs.

In essence, an ABM is built up by a collection of entities called *agents*, whose state and behavior are governed by a set of computational *rules*. Agents “live” in the virtual *world,* which is discretized into grids, or *patches*. Each *patch* is a certain situation or a physical environment that an agent lives in. In the application of social behaviors, each agent can represent an individual person with distinctive features with respect to demography, psychology and social history. Second, an individual agent makes its own decision and adapts the behavior to the environment over time that effectively represents the dynamics of social and physical influences on human behavior in the real world [43]. Lastly, ABM is technically flexible for users to explore a wide parameter space and a long temporal scale, which otherwise would be very costly or at times prohibitive with empirical experiments. For example, ABM can simulate human interactions in social networks with the ability to control for network sizes, type and frequency of interaction, and observe individual and group outcomes from weeks to years. Given that the process of stigmatization is multiscale and time-evolving, ABM presents a new, exciting opportunity in the research of health stigma.

### Study Objectives

The objective of this study was to investigate the experience of vocal stigma using empirical survey questionnaires and computer network models. The primary focus was to predict how peer interactions could affect an individual’s help-seeking behaviors and vocal stigma experience especially in vocal performers. With a better understanding of vocal stigma, clinicians, social epidemiologists and public health policy makers will be more informed of devising interventions aimed at reducing vocal stigma and improving access to voice healthcare.

## 2. Methods

### 2.1. Empirical Study of Vocal Stigma Profiling

#### 2.1.1. Survey Development and Deployment

The study protocol (A09-B73-20A) was approved by the Institutional Review Board at McGill University, Canada. A 64-item questionnaire was constructed to survey individuals’ demographics (6 items), occupation and training (3 items), vocal health history (14 items), IMB profile (30 items, 10 each for information, motivation, and behavioral skills), experiences of vocal stigma (10 items, 5 each for social-stigma and self-stigma) plus a single, open item for feedback. The questionnaire was developed in consultation with an expert panel, including: (a) a speech-language pathologist with over 10 years of experience working with clinical voice disorders; and (b) three professional vocal performers (one actor, one singer, one actor/singer; each with over 20 years of experience) from the Alliance of Canadian Cinema, Television and Radio Artists (ACTRA) or the National Association of Teachers of Singing (NATS). Specific details of the survey construct and coding scheme are provided in ***Supplementary Information: Additional File 1***. The survey was deployed via *LimeSurvey*, Version 3 (LimeSurvey GmbH, Hamburg, Germany), an open-source survey tool housed at McGill University. The survey took participants around 10-15 minutes to complete all items.

#### 2.1.2. Participant Recruitment and Compensation

A convenience sample of 200 professional vocal performers and 203 gender-matched controls were recruited for this study. Professional vocal performers were recruited through membership offices of ACTRA and NATS. For this study, professional vocal performers were defined as those receiving at least part of their income via singing or acting performance. For the gender-matched control group, an online recruiting platform Prolific (www.prolific.co) was used for recruiting non-vocal performers to the study. Non-vocal performers were defined as receiving no income via singing or acting performance and having no past or present employment in the arts sector listed in their participant profiles on Prolific.

Participants without a valid Canadian postal code were excluded from this study because an individual’s stigma experience and tendency to seek help are specific to their own countries’ healthcare systems. Participants were also excluded if they had a history of vocal pathology arising from cancer, stroke, degenerative neurological conditions, or physical trauma to the throat, head and neck.

Interested individuals would first need to answer a set of screening questions to ensure their eligibility on *LimeSurvey*. Eligible participants were directed to the consent form, in which the general purpose of the study was described. Explicit reference to stigma was omitted to avoid response biases. A debrief with the full purpose of the study was disclosed to participants at the end of the questionnaire, at which time they could opt to withdraw their data from the study. In this study, no opt outs were requested at the end.

In terms of compensation, individuals in the vocal performer group, were given the choice to enter a raffle for gift cards (each CAD $50) by following an external link and providing their email address at the end of the study. Participants in the control group were paid CAD $5 via the Prolific platform.

#### 2.1.3. Data Privacy

All data were collected anonymously and were stored on a secure server, hosted by McGill University. Participant data did not contain any identifying information. The emails in the gift card raffle were not linked to individual participant data. Participant identity was thus not traceable or identifiable. No additional information was collected passively about participants’ computers, e.g., IP address, cookies etc.

#### 2.1.4. Preparatory Data Analysis

Given that the survey data is the data source for ABM development, we opted to summarize the statistics of the empirical data here to avoid duplication and confusion from those of simulation experiments in Section 3. In short, a total of 200 professional singers and actors and 203 controls participated in the survey (both groups: ages 21-65; female 65%, male 32%, other 3%). Full details of the empirical data, participant demographics and survey data statistics can be found in ***Supplementary Information: Additional File 2, Table 2.1-2.3*** and ***Additional File 3, Table 3.1-3.6*** respectively.

With respect to vocal stigma experience, performers reported 14% more stigma than controls (*t*(401)=8.87, *p*=0.025) overall. Among professionals, singers reported 15% more stigma than actors (*t*(198)=-1.67, *p=*0.025). Irrespective of groups, stigma was negatively associated with (1) *age* (Performers: *r*=-0.27, p<0.001; Controls: *r*=-0.17, *p*=0.018), (2) *recency of a voice disorder* (Performers: ρ=0.15, *p*=0.033; Controls, ρ=0.14, *p*=0.047), and (3) *frequency of voice disorder* (Performers: ρ=0.46, *p*=0.005; Controls: ρ=0.26, *p*=0.031).

To assess the association of stigma experience with IMB profiles, Pearson’s product-moment correlations showed that stigma correlated negatively with motivation (Performers: *r*=-0.49; Controls: *r*=-0.59, *p*<0.001) and behavioral skills (Performers: *r*=-0.28; Controls: *r*=-0.46, *p*<0.001) in both participant groups. Stigma was not significantly associated with information (Performers: *r*=-0.09, *p*=0.205; Controls: r=-0.09, *p*=0.200), meaning there was not a definitive trend between stigma and an individual’s knowledge/beliefs regarding voice disorder or vocal health.

Broadly speaking, vocal stigma is found present among professional singers and actors in Canada. The level of Motivation and Behavioral Skills of an individual were negatively associated with vocal stigma. The negative association between age and experiences of stigma may indicate that early-career vocal performers are more vulnerable to this stigma. The positive association between a history of vocal illness and experience of vocal stigma may indicate that vocal stigma is not commonly recognized by individuals without direct experience with a voice condition.

### 2.2. Computational Study of Simulating Social Interactions and Vocal Stigma

#### 2.2.1. Vocal Stigma Agent-Based Models: Conceptual Framework

In this vocal stigma agent-based model (VS-ABM), each agent represents an avatar of a vocal performer (VP) or a non-vocal performer (non-VP), who functions as a social actor and interacts with their peers to share information and their motivations for help-seeking related to voice health. Depending on an agent’s geolocation, they have access to a small or a large peer network in the VS-ABM. When two agents interact, they will exchange information and motivations related to vocal health.

An agent’s vocal health (history and frequency of disordered voice), stigma experience (self- and social-stigma) and IMB are connected via two feedback loops (social and personal), which in turn affects an agent’s tendency to seek help **(Figure 3)**. The social loop simulates the interrelationship between social-stigma and IMB. Also, an agent’s motivation is linked to its own frequency of voice disorder. For instance, if an individual has frequent relapses in voice disorders, they will likely have less motivation to seek help from social partners and professionals. In contrast, the personal loop simulates the interrelationship between self-stigma and IMB. Also, an agent’s information related to vocal health is contingent on its history of voice disorder. For instance, if an individual has prior experience with a voice disorder, they will likely have more information about the nature of it and may have reduced their self-stigma as a result.

**Figure 3.**
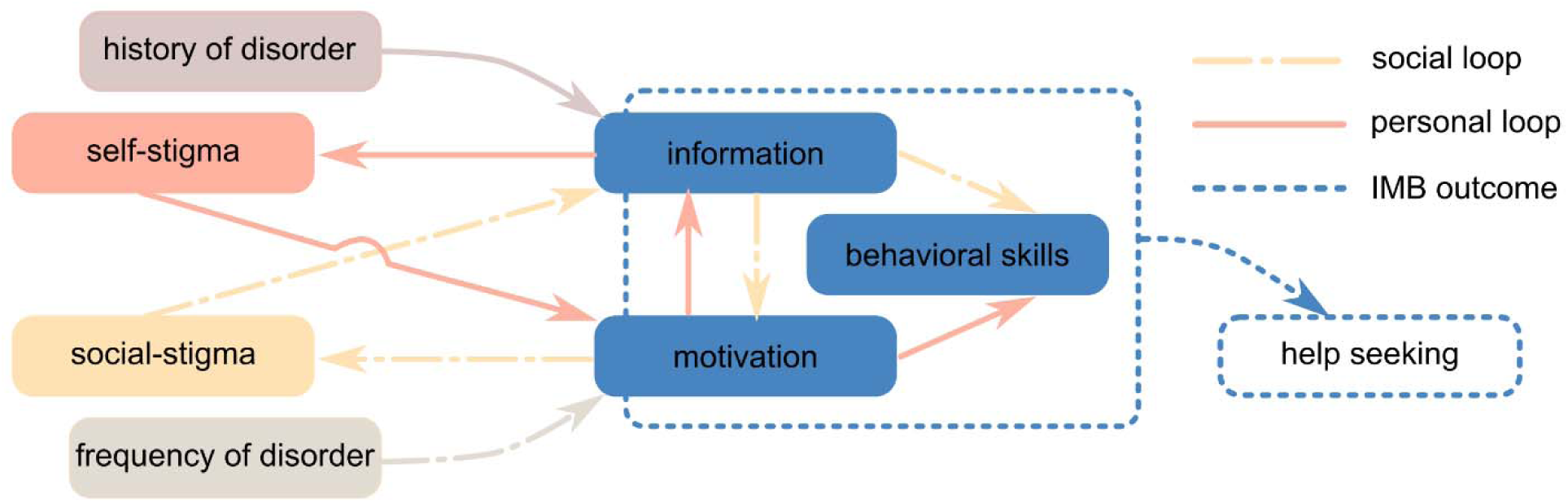
The Framework of Vocal Stigma Agent-Based Models (VS-ABM). An agent’s vocal health, vocal stigma, information, motivation, and behavioral skills (IMB) are presumably interrelated and modulated via the social and personal feedback loops, which collectively affects an agent’s tendency to seek help.

#### 2.2.2 VS-ABM Design: World

The VS-ABM is developed with NetLogo version 6.3.0, a free and open-source, cross-platform, agent-based modelling framework [67]. In our implementation, the VS-ABM world represents a small urban community, which is divided into a total of 1089 patches (33 x 33), with a densely populated urban center and a sparser rural perimeter. This geographical specification of the VS-ABM is based on the following considerations. (**Table 1**)

**Table 1.**
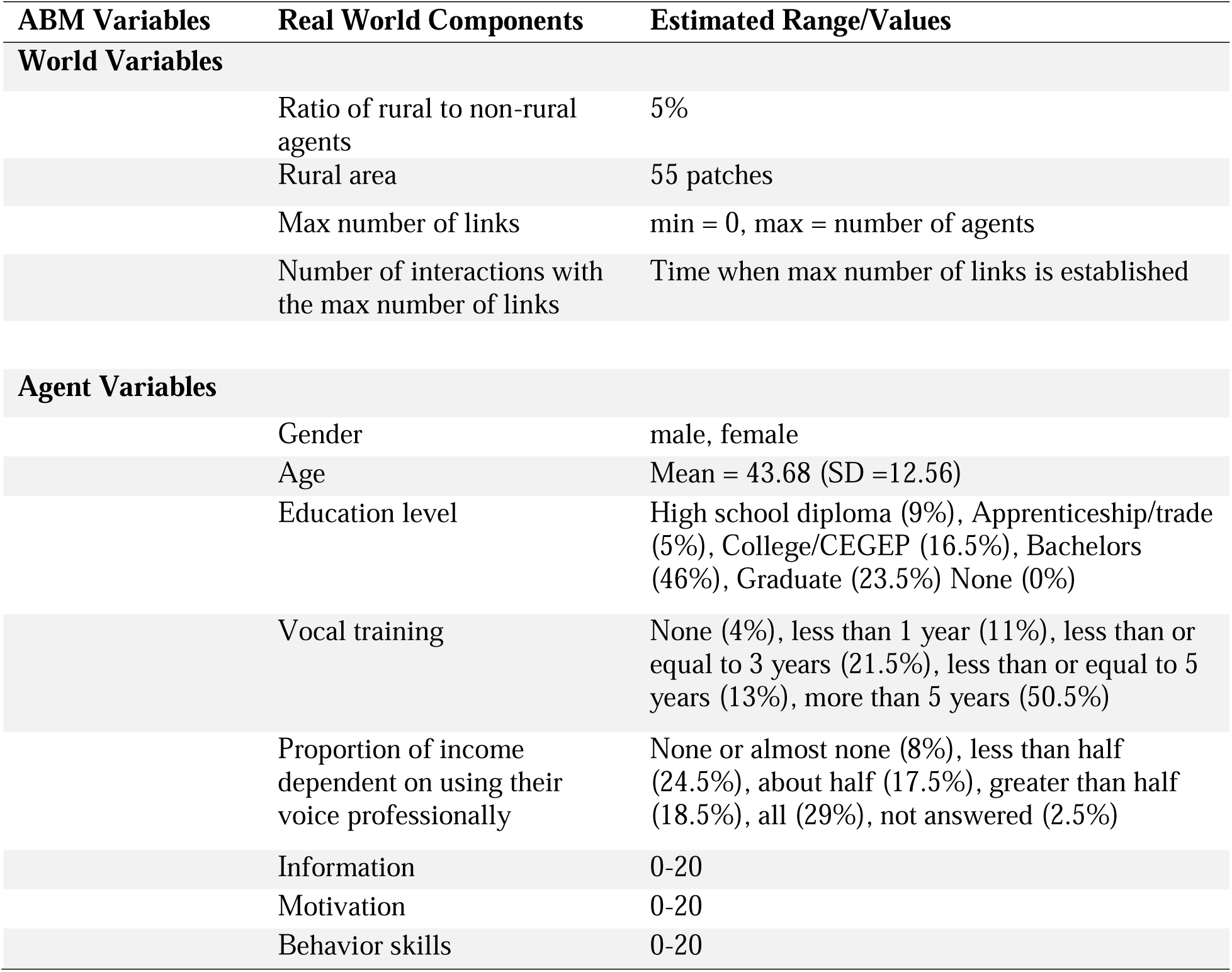
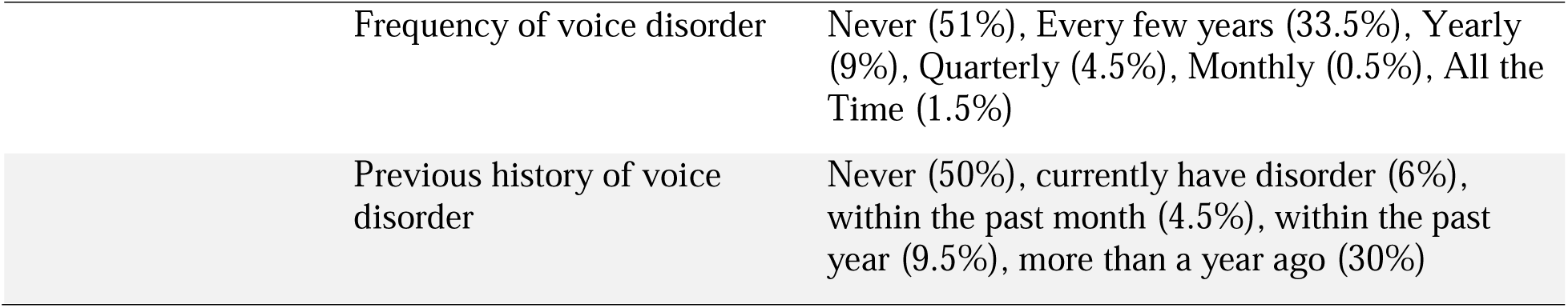
Variables of World and Agents in the Vocal Stigma Agent-Based Model.

| ABM Variables | Real World Components | Estimated Range/Values |
| --- | --- | --- |
| **World Variables** |  |  |
|  | Ratio of rural to non-rural agents | 5% |
|  | Rural area | 55 patches |
|  | Max number of links | min = 0, max = number of agents |
|  | Number of interactions with the max number of links | Time when max number of links is established |
| **Agent Variables** |  |  |
|  | Gender | male, female |
|  | Age | Mean = 43.68 (SD =12.56) |
|  | Education level | High school diploma (9%), Apprenticeship/trade (5%), College/CEGEP (16.5%), Bachelors (46%), Graduate (23.5%) None (0%) |
|  | Vocal training | None (4%), less than 1 year (11%), less than or equal to 3 years (21.5%), less than or equal to 5 years (13%), more than 5 years (50.5%) |
|  | Proportion of income dependent on using their voice professionally | None or almost none (8%), less than half (24.5%), about half (17.5%), greater than half (18.5%), all (29%), not answered (2.5%) |
|  | Information | 0-20 |
|  | Motivation | 0-20 |
|  | Behavior skills | 0-20 |

|  |  |
| --- | --- |
| Frequency of voice disorder | Never (51%), Every few years (33.5%), Yearly (9%), Quarterly (4.5%), Monthly (0.5%), All the Time (1.5%) |
| Previous history of voice disorder | Never (50%), currently have disorder (6%), within the past month (4.5%), within the past year (9.5%), more than a year ago (30%) |

First, the VS-ABM world size was set to approximate to the lower boundary of a sparse geographi network. This is because simulated social network in all sizes need to fit within the ABM world without being too densely populated. If the ABM world is too big, agents will be too far away from each other for social interactions. Second, the VS-ABM world is set as box-bound. That is, agents on the left or the top edges do not interact with the agent on the right or the bottom edges and vice versa. That way, agents in the rural region at the outer border are constrained to have fewer possible connections compared to those in the urban center of the simulation space. Note that about 5% of survey respondents were from rural areas in Canada (***Supplementary Information: Additional File 2, Table 2.1*)**. To approximate this real-world geographical distribution in the VS-ABM, we set the smallest network (50 agents) to have around 5% patches occupied by agents, whilst the same proportion of rural agents approaches 100% of occupied patches for the largest social network (400 agents). This geographical specification allows the proportion of rural agents to remain 5% in both geographically sparse networks (with 50 agents) and socially sparse networks (with 400 agents).

#### 2.2.2 VS-ABM Design: Individual Agent Attributes

Each agent has a set of attributes in terms of personal background (e.g., gender, age, education level, vocal training etc.) as well as experience related to voice health (e.g., information, motivation, behavior skills, self-stigma, social-stigma, history of voice disorder). (**Table 1**) All attributes are randomly assigned to each agent based on the normal distribution of the values from the demographic data in the empirical study. (***Supplementary Information: Additional File 2)***

#### 2.2.3 VS-ABM: Agent-Agent Social Interactions

Within the ABM world, agents are free to interact and exchange information with nearby peers. These agents cannot move their location and are surrounded by neighbouring peers whom they are free to interact with. Agents also randomly select their next agent for each interaction. The potential peers are limited to a circular region around each agent with a radius of three patches. In other words, agents’ interaction space is 2.5% of the possible geographic space. We also assume that an agent with high initial social-stigma may be reluctant to interact with their peers. As such, in the VS-ABM, if either agent has high social-stigma (over 7.5), the chance of their peer interaction is set as 75%.

After each interaction, the IMB and vocal stigma of an agent will change incrementally. The exact incrementation is formulated based on the linear regression analyses of help-seeking and stigma relationship in the survey study (***Supplementary Information: Additional File 3, Table 5)***. For example, within the social loop, social-stigma is associated with information. After each peer interaction, an agent’s information will slightly change by a value that is randomly sampled from a normal distribution centered around the mean, as obtained from the corresponding regression analysis (Information-Social-Stigma: 0.066). The change in an agent’s information can also cascade to changes in behavior skills and motivation. The formulation is the same but with different center means derived from the corresponding regression terms (Information-Behavioral Skills: 0.232; Motivation-Social-Stigma: 0.316). The formulation of all agent-rules is detailed in ***Supplementary Information: Additional File 4, Table 4.2***. Below is pseudocode for the VS-ABM.

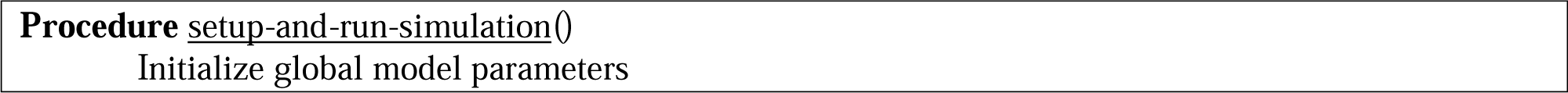

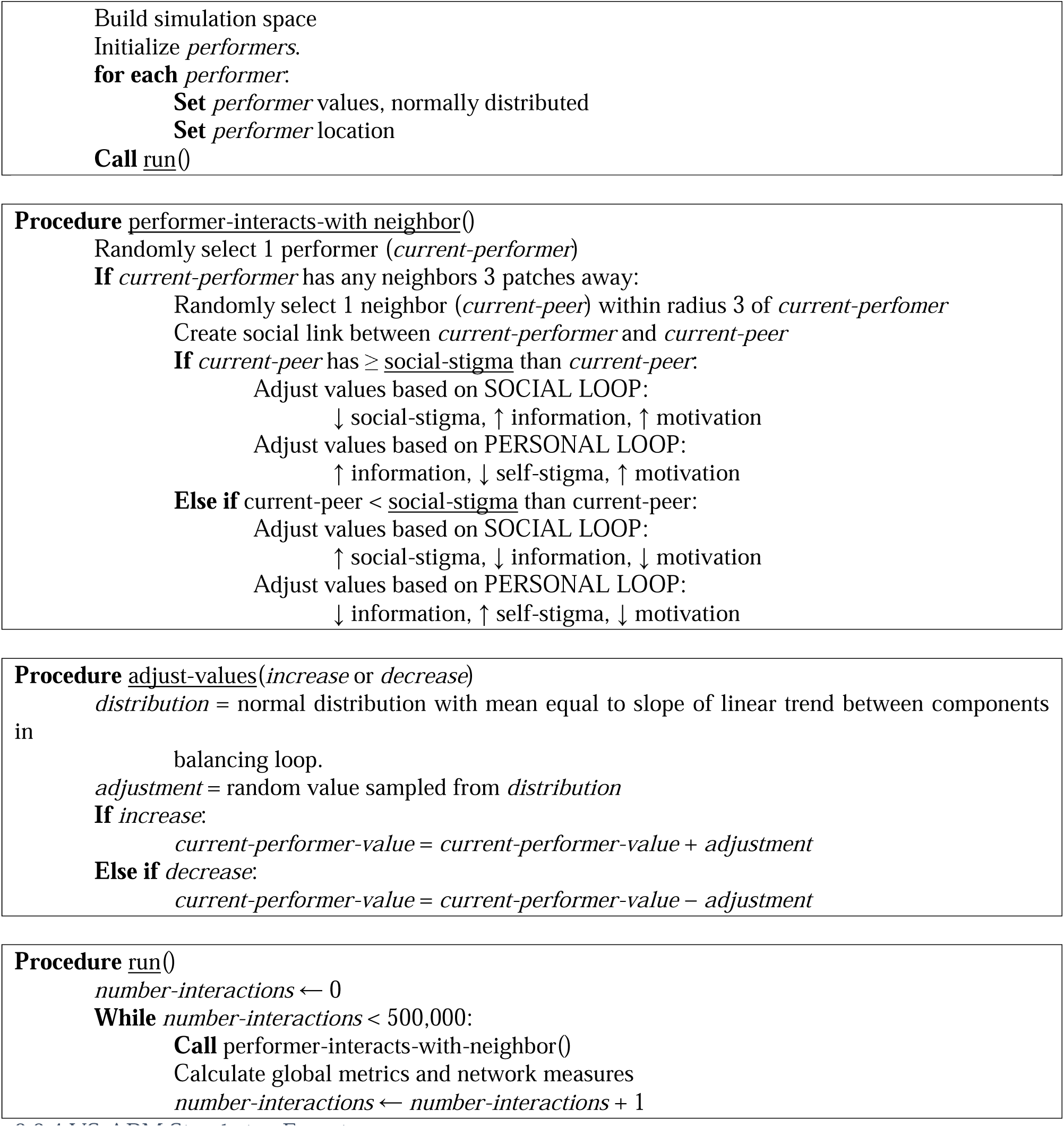

#### 2.2.4 VS-ABM Simulation Experiments

This VS-ABM is designed to simulate social interaction and transfer of information/motivation among peers as functions of network size and initial social-stigma. Simulation experiments were set up to answer three research questions respectively *(**Supplementary Information: Additional File 4,*** ***Table 4.1 for the ABM world initialization and Table 4.3 for the initialization procedure of simulation experiments***).

##### Research Question #1: Does vocal stigma decrease when there are larger social groups?

To investigate the relation between network size and total stigma, we ran 500 simulations for each network size with 50, 100, 150, 200, 250, 300, 350 and 400 agents up to 500,000 social interactions. We hypothesize that an agent’s total stigma will reduce with increasing peer networks. With more peers available to interact with, agents have a greater variety of peers to gain experiences and information from. More peers, more information, and more interactions should make agents more likely to increase their total IMB and lower their vocal stigma. Peers with extreme values for IMB and stigma may polarize their limited network more severely than with a larger peer group.

##### Research Question #2: Does vocal stigma decrease when peers are more willing to socially interact?

To evaluate social willingness for agent-agent interactions, we ran additional 800 simulations initialized for five agent populations with distinctive initial social-stigma profiles, since empirical data showed differences between non-VP and VP groups *(**Supplementary Information: Additional File 3,*** ***Table 3.1-3.4)***. Each group is assigned with an initial social-stigma value obtained from the empirical data *(**Supplementary Information: Additional File 3,*** ***Table 3.6)***. For instance, the initial social-stigma value for the “General Population” is 5.4, which is derived by averaging social-stigma scores from non-VPs and VPs. While the VS-ABM does not explicitly model medical intervention, we assume that social interventions based on peer interactions will act similarly on social-stigma. Related hypotheses are: (1) social-stigma will decrease over time for agents who are more willing to interact; and (2) agents with high social-stigma will be less likely to transfer information/motivation to peers. A total of 4000 simulations were run for the following five agent populations.

1. General Population of both non-VPs and VPs with average social-stigma and average risk and incentive.
2. non-VP group with low social-stigma with low risk of voice disorder and little incentive to seek professional help.
3. VP group with higher social-stigma with higher risk of voice disorder and more incentive to seek professional help.
4. VP group with a history of voice disorder having very high social-stigma elevated in part by no intervention for the voice disorder.
5. VP group with a history of voice disorder having high social-stigma that has been mitigated by some social intervention.

##### Research Question #3: How does social network structure impact IMB and stigma?

A comprehensive network-based analysis was employed to evaluate the impact of social network structure on the flow of information and motivation among agents for all 4000 simulations run. A NetLogo network extension procedure (*“nw”*) was used to quantify two standard network measures, namely, the degree of a node and local clustering coefficient [68]. The degree of the node (*count links*) is used to measure the number of peers that an agent is connected to in the network. The local clustering coefficient (*nw:clustering-coefficient*) is used to measure how densely connected an agent’s neighbors are and how much they tend to group together in a network by averaging all individual nodes. While this analysis could be performed on both the network size and social-stigma manipulations, we chose to focus on network size for two reasons. First, social structure in these networks should be directly correlated to the number agents in the simulation, since the simulation space does not change nor does the radius which agents survey for neighbors. Second, in order to generalize across all simulations, we use an outcome measure agnostic to initialized values of social-stigma, change in total stigma, which also allows us to compare directly with change in IMB so they are on similar measurement scales. We thus test which network structure, degree (number of peer connections) or clustering coefficient (peer connectedness), has the largest impact on change over time.

## 3. Results

### 3.1 Overall Relation between Total Stigma and Social Network Size

Our original hypothesis for Research Question #1 was that vocal stigma would decrease with larger social networks. Unexpectedly, this hypothesis was not supported by our ABM simulation results in looking at changes in total stigma relative to the number of interactions with network sizes (50-400 agents) and initial social-stigma profiles (low vs average vs high) for each VP, non-VP and general populations (**Figure 4A**).

**Figure 4.**
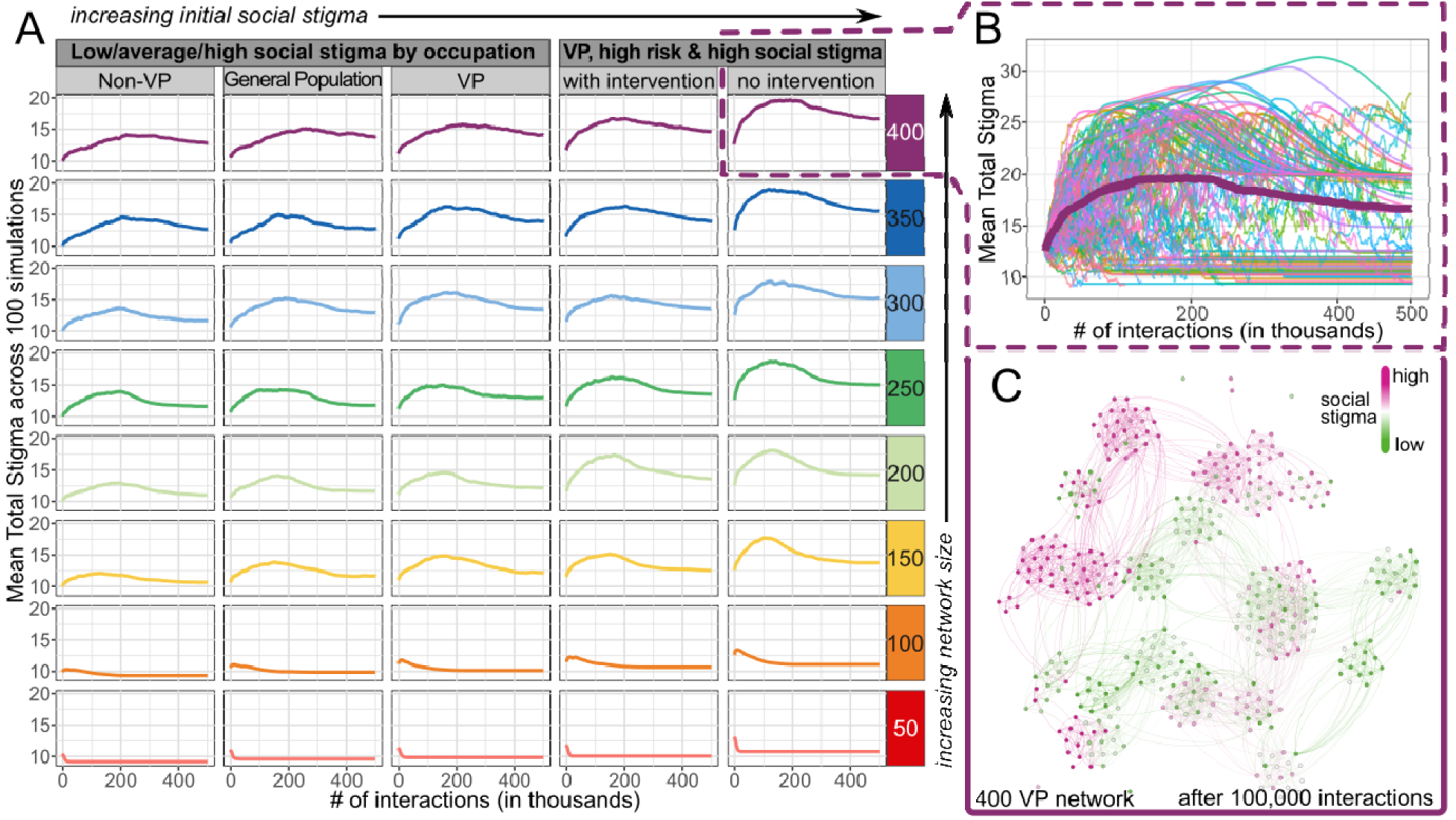
Total Stigma Experience as Functions of Network Size and Number of Interactions in Agents with Low, Average and High Initial Social-Stigma. (A) Aggregated trends of mean total stigma scores. Each trend aggregated from 100 simulations initialized with social-stigma and network size. (B) Varying outcomes of mean total stigma score from individual simulations for VP agents with high risk of voice disorders, high initial social-stigma, and no intervention within the 400-agent network size. The aggregated trend, same as VP group in Fig 4A, for all 100 simulations is shown in bold. (C) A representative snapshot of simulated social networks with 400 VP agents near the maximal total stigma after 100,000 interactions in the 400-VP network. An agent’s social-stigma is color-coded from low (Green) to high (Red).

First on the VP networks, agents in the two smallest network sizes (50 and 100) had their total stigma stabilized as early as 25,000 social interactions. Their final total stigma values were also lower than initial values, regardless of initial social-stigma. In contrast, when the networks became sizable (> 150 agents), the trajectories of total stigma started to follow a log-normal distribution pattern. That is, agents’ total stigma quickly increased to a peak and then slowly leveled off but did not always return to initial values. Interestingly, VP agents with high initial social-stigma took 25% fewer social interactions (50,000 to 100,000) to reach the peak of total stigma, compared to those with low initial social-stigma (the general and non-VP groups). This initial observation suggests that total stigma likely worsens over time with social interactions in larger network groups, particularly for individuals prone to higher social-stigma such as VP at high risk for voice disorder and a tendency to not seek intervention.

To further examine this paradox, we proceeded to examine individual-specific variations in the evolution of total stigma in the largest network (400 agents) from the VP group with initial high stigma up to 500,000 interactions (**Figure 4B**). Three distinct patterns emerged from this case simulation. (1) First, one set of simulations quickly leveled off towards a total stigma value lower than the initial value, which is denoted as the “*decreasing cohort*”. (2) Another set of simulations, which is denoted as the “*increasing cohort*”, increased rapidly towards a maximum between 50,000 and 200,000 interactions and then slowly leveled off to a total stigma value larger than the initial value. (3) Lastly, the “*chaotic cohort*” is a set of agent simulations that appeared to fluctuate randomly and did not appear to stabilize even after 500,000 social interactions. An extended set of simulations were further run for the “*chaotic cohort*” and confirmed that the total stigma value continued fluctuating randomly up to 10,000,000 interactions (data not shown).

Except for the “*chaotic cohort*”, most agents’ total stigma appeared to approach maximum around 100,000 social interactions with large social networks. To understand one such maxima, we further visualized an established social network from the “*increasing cohort*” in 400 VP agents after 100,000 social interactions (**Figure 4C**). Multiple clusters or social groups with similar social-stigma values were formed within the global network, suggesting a homogenization with these highly interconnected social groups. Also, these clusters were not isolated but rather fairly connected to neighboring groups. In other words, peers were connecting with the whole set of the neighboring group, rather than simply bottlenecking through a few nearby peers.

In particular, a few social groups appeared to have a heterogeneous mix of agents with initial low and initial high social-stigma profiles (**Figure 4C: networks with mixed green and red color**), confirming that agents could interact with opposite social-stigma values. Especially in this “*increasing cohort*”, the emergence of mixed social groups in the global network coincides with the stabilization of total stigma around the critical point of 100,000 social interactions. We speculated that, at the beginning of interactions, agents would establish social groups with peers around them, becoming more like their neighbors. Then, when social groups became homogenous, the total stigma would approach to its peak. Eventually, when social groups with polarized views interacted, a mix of agents with low and high social-stigma emerged and, paradoxically, stabilized the population’s social-stigma.

To answer Research Question #1, our simulation results indicated that, on average, vocal stigma increases with larger social groups. However, there are non-trivial trends where total stigma diminishes as social groups become less polarized. In addition, our results suggested that interactions between groups are not bottlenecked with a small set of agents but distributed among the entire social group. The clusters can exchange information/motivation with multiple members of neighboring groups, which may ultimately result in less polarized neighboring groups.

### 3.2 Social Network Size in relation to Each IMB and Vocal Stigma Components

For Research Question #2, the original hypothesis was that social-stigma would decrease when agents were more willing to socially interact, i.e., lower social-stigma and higher IMB. This hypothesis was partially supported by our ABM simulation results in comparing the probability distributions of final IMB and final total stigma scores between the smallest and largest social networks (50 vs 400 agents) (**Figure 5A-B**).

**Figure 5.**
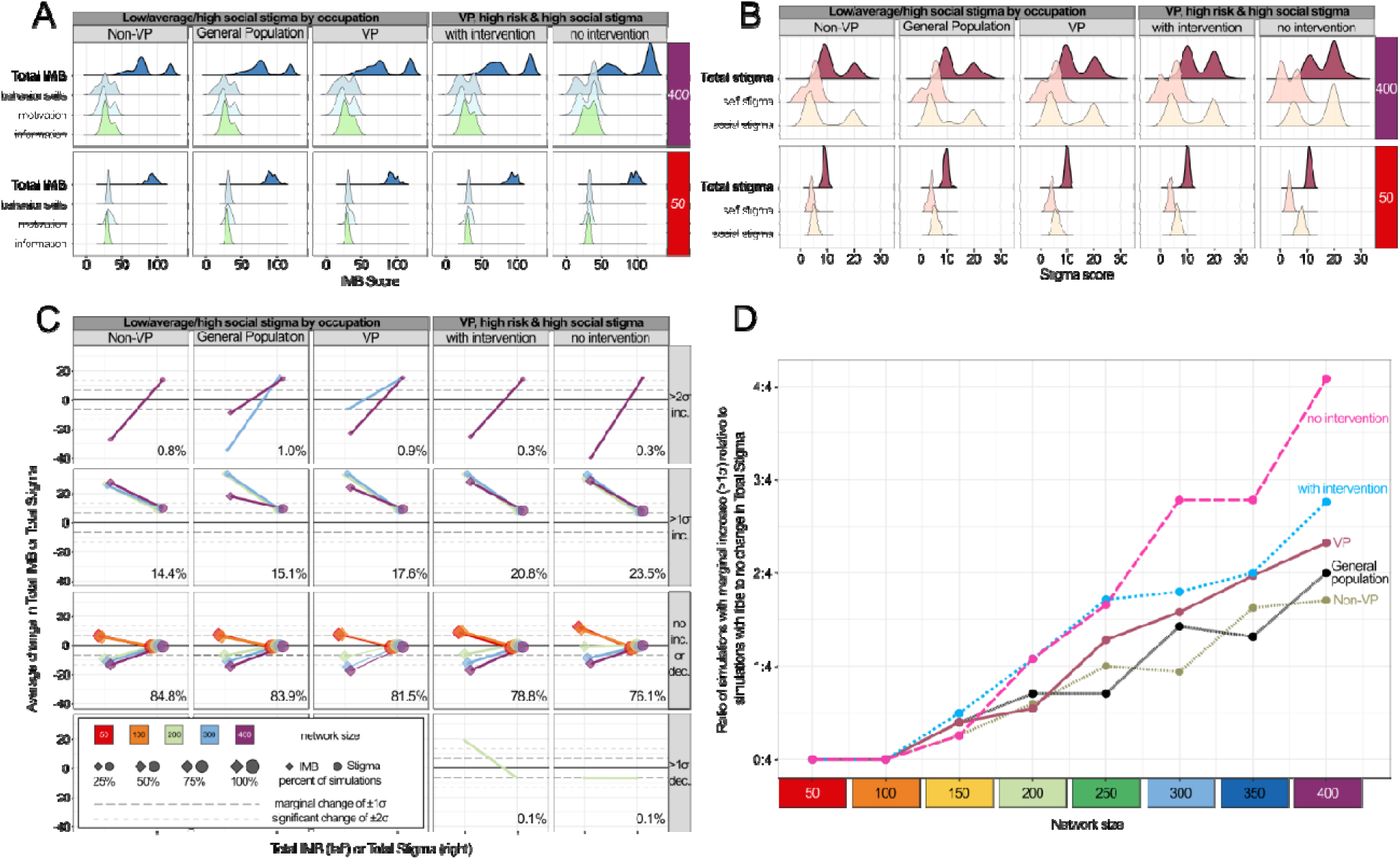
Probability Distribution of Total IMB and Total Stigma Experience. A total of 500,000 social interactions were simulated for VP, Non-VP and general populations with low, average or high initial social-stigma. (A) Total IMB and its components (information, motivation, and behavioral skills) at 50 and 400 network sizes (B) Total stigma and its components (self-stigma and social-stigma) at 50 and 400 network sizes (C) Average change in total IMB (diamonds) and total stigma (circles) from their initial values. Each row of subplots represents a specific magnitude of change, i.e., from less than 1 SD (σ) to more than 2 SD increase (inc.) or decrease (dec.), in total IMB and total stigma. % = percentage of simulations with corresponding changes in total stigma across all network sizes. (D) Ratio of simulation with total stigma changes more than 1 SD (Panel C row 1 + 2) relative to those with little or no change (Panel C row 3).

Following a total of 500,000 social interactions, a clear bimodal distribution of total IMB score was noted in large but not in small networks across all three agent populations (**Figure 5A**). Of the total IMB score, the first mode (local maximum) appeared around 60 and 75, whereas the second mode appeared around 110 in the probability density function. Each IMB element (information, motivation and behavioral skills) also appeared to be highly coupled. That is, an agent population, regardless of its initial social-stigma, can be sub-grouped into either with low IMB and high IMB. Such sub-group distinction is much more pronounced in large social networks than small ones.

For the total stigma, a bimodal distribution also appeared in large but not small social networks across all three agent populations (**Figure 5B**). Of the total stigma score, the first mode was centered around 10, whereas the second mode was around 20 within the data. Each total stigma element (social- and self-stigma) also displayed two distinctive peaks, but their distance was more separated in social-stigma score. Additionally, comparing high-risk VP groups, those with social intervention had overall lower social-stigma, whereas the maximal mode for the no intervention group showed higher social-stigma. Our results indicated that social-stigma experience has more weight than self-stigma in the total stigma experience.

Further analysis was performed to evaluate the magnitude of changes in total IMB and total stigma by aggregating VP, non-VP, and general populations before and after the 500,000 interactions (**Figure 5C-D**). For total IMB, about 35% and 63% of agents showed increases or no changes of their scores in smaller networks (50 and 100 agents) at the end of simulations. With larger network sizes (>150 agents), about 36% of agents showed significant increases in their total IMB scores with more than two standard deviations (SD, σ), i.e., between 30 and 40 points increase from initial values. At the same time, about 39% and 21% of agents showed reduction in their IMB scores more than 1 and 2 SD respectively in these larger networks (**Figure 5C**). For total stigma, all agents in smaller networks had negligible changes of their scores, whilst 25% of them had total stigma increased by more than 6 points when they were in larger networks. For those with high risk and high social-stigma, VP agents were 1.5 times more likely to experience more total stigma if they had no intervention, compared to those with social intervention, when they are in the largest social network (**Figure 5D**).

All in all, the aforesaid simulation results confirmed that higher total IMB was associated with lower total stigma for small networks. Interestingly, IMB outcomes vary across network sizes. Despite minimal change to total stigma, IMB tended to decrease more for larger social networks, while IMB for smaller networks tended to increase. The bimodal distribution noted in large social networks (i.e., 400) may be attributed to the increased likelihood that extreme values of social-stigma arise in the population. Also, within a large social network, their social groups tend to be more polarized with group size as noted in **Figure 4C**, leading to a wider separation between the two modes of the bimodal distributions.

To address Research Question #2, our results supported the hypothesis in cases of small networks, i.e., lower social-stigma leads to an overall decrease in total stigma. Larger networks are far more likely to have significantly increased total stigma. For high-risk VPs, social interventions might help mitigate high social-stigma and eventually a reduction in total stigma over time.

### 3.3 Network Structure in relation to Total IMB and Total Stigma

To understand the connections among agents in the social network, degree of node and clustering coefficient were computed in relation to the changes in total IMB and total stigma when the social network was established (i.e., after 100,000 social interactions) in all 4000 simulations.

First, the degree of node represents the number of social links that an agent has in a social network. (**Figure 6A**) The average number of links ranged from 2.5 in the smallest social network (50 agents) to 13.5 in the largest social network (400 agents). This positive linear relationship is expected because each agent has more peers to interact within their vicinity in larger social networks. For instance, the relative patch coverage increases from 5% in the 50-agent network to nearly 36% in the 400-agent network.

**Figure 6.**
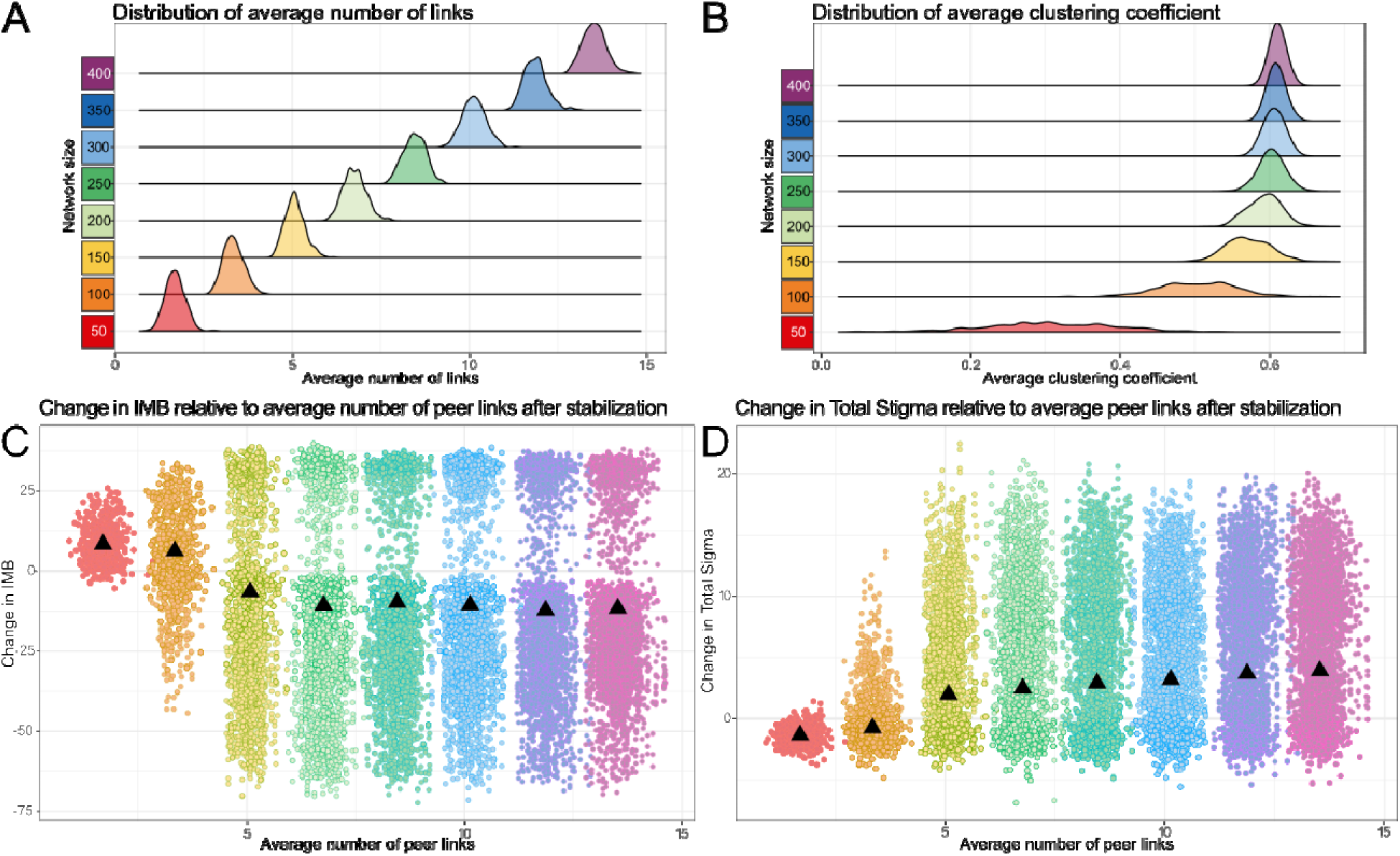
Network Structure Analysis of Network Size Effects on IMB and Vocal Stigma. (A) Distribution of average number of peer links (degree of nodes) by network size. (B) Distribution of average local clustering coefficient by network size. (C) Total IMB values decrease as a function of average number of peer links, which is highly correlated with network size. A clear bifurcation appears around 4.5 peer links. (D) Total stigma values increase as a function of average number of peer links. No bifurcation appears. However, distributions show a larger range of changing stigma values for more peer links.

Second, the clustering coefficient represents how close-knit an agent’s social group (i.e., cluster) i interconnected in a social network. The clustering coefficient is measured on a scale of 0 to 1. A lower cluster coefficient is when peers are only connected through the agent, whereas a higher cluster coefficient is when agents’ peers connected independently and create denser and tighter connections.

Interestingly, the average clustering coefficient for the network does not show a linear trend with network size (**Figure 6B**). A wide spread of clustering coefficients was observed in small social networks wherea the probability distribution approximated to a normal distribution towards a larger social network. The cluster coefficient is also higher in larger networks (400 agents; mean (SD) = 0.61 (0.009)) than that of smaller networks (50 agents; mean (SD) = 0.31 (0.086)). In small networks, agents were sparsely randomized in the simulation space, which geographically favored agents being connected with their neighboring peers only but not reaching out to other social groups that are further away.

Finally, we analyzed how the number of peer links (i.e., social partners) affects an agent’s total IMB and total stigma scores when the social network is stabilized, i.e., all links are maximally connected. For total IMB, more peer links might relate to a gradual decrease in IMB. (**Figure 6C**) Upon the stabilization of social networks, the IMB increases by 7.6 points in the smallest 50-agent network (1.5 social links); whereas it decreases by 10.6 points in the largest 400-agent network (12.5 social links). For total stigma, more peer links might contribute to a slight increase in total stigma (**Figure 6D**). Upon network stabilization, the total stigma decreases by 1.2 points in the 50-agent network yet increases by 3.5 points in the 400-agent network.

## 4. Discussion

By using a prospective survey and network simulations, we were able to: (1) characterize population behavior related to vocal stigma in professional vocal performers; (2) identify which IMB and stigma components contribute most to population outcomes; and (3) analyze network structure and social groups to understand the impact of social interactions on population outcomes. Our VS-ABM allows each IMB and stigma component to be examined independently while simultaneously modelling how each element influences others. These components can vary for each individual and change as individuals interact with peers with distinct IMB and health profiles.

### 4.1. Diverging population outcomes and heterogeneity within social groups

Three population behaviors, namely decreasing, increasing, and chaotic cohorts, were identified across all network sizes. The “*decreasing cohort*” had characteristics that resembled the small network trends, such that total stigma decreases and quickly stabilizes at a value less than the original population stigma. The “*increasing cohort*” had characteristics that resembled the large network trends, where total stigma increases to a maximum and then levels off towards a value larger than the initial stigma value. Such stigma maxima are seen when the social network of the population is fully established, and social groups are largely homogenous. When polarized homogenous groups start to interact and exert more influence on each other, greater heterogeneity of the groups emerges at a population level. Lastly, the “*chaotic set*” is referred to an oscillatory behavior when total stigma does not stabilize. Such complex patterns of vocal stigma are not programmed (i.e., hard-coded) in VF-ABM but rather emerge *de novo* (i.e., emergency) from dynamical systems. Our simulation outcomes may also be in line with a new notion of “stigma mutation”. [69] For instance, stigma associated with COVID-19 has a social-historical context and changes over time in response to the local and global environment. In our case, agents’ social-personal backgrounds (e.g., occupation, medical history, initial social-stigma) modulate the evolution of their total stigma in a network environment.

### 4.2 Social-stigma drives change in vocal stigma

We further investigated which specific IMB and stigma components drove changes in population outcomes in relation to network size. Overall, information and behavioral skills tend to be coupled more strongly with motivation being inversely related, i.e., higher information and behavioral skills relative to a lower motivation. Social-stigma has more impact than self-stigma on total stigma. The variance of IMB changes is considerably large across all network sizes, ranging from −6 to +5 SD changes on average. In contrast, the variance of total stigma changes remains quite consistent – around 75%-85% of all simulations show less than 1 SD change on average. Moreover, in larger network sizes, social interactions led to three distinctive patterns of social-stigma, namely, (1) small decreases in low/ average risk groups, (2) small increases in high-risk groups; or (3) large increases regardless of risk group. In particular, larger networks with higher risk groups of voice disorders have considerably higher odds of increasing the total stigma than those of lower risk groups. Among these higher risk individuals, those without medical interventions have higher odds (4:4) to increase total stigma after social interaction than those who received treatment (3:4). That is, for large networks (i.e., 400 agents), about 51% of VP with no intervention are likely to increase their stigma after interacting with their peers; this likelihood drops to 41% if they receive intervention.

Another key observation is that IMB and stigma do not always have a directional relationship. For instance, if we looked at simulated populations whose total stigma changed more than 1 SD with social interactions, their IMB scores could increase by 20 points at most. For those with changes more than 2 SD in total stigma, IMB decreased significantly by 30 points – in this case, vocal stigma might get worse despite informational and social intervention. Similar notions were also confirmed in reducing public stigma of mental illness, in which stigma is difficult to eliminate especially if it is deep rooted in social relationships [70].

### 4.3 Larger social groups may promote vocal stigma

Two standard network measures were analyzed to evaluate how connected VPs were (i.e., degree of node) and how close their social groups were linked (i.e., clustering coefficient). As the network size increased, a VP agent had more peers to interact and connect with in their vicinity. That is, for larger networks, peers had consistently larger social groups. However, as we simulated hundreds of thousands of social interactions, all agents were able to eventually interact and connect with all their potential peers and all social groups were maximally interconnected or clustered. As social group networks stabilize and become maximally connected, a gradual increase in total stigma and decrease in IMB was evident in larger but not smaller social groups. These analytics also provided part of the evidence to answer Research Question #1, i.e., larger networks stabilized towards more polarized social groups. This observation aligns with similar social network findings of polarization of political groups and how negativity spreads through larger social networks faster than positivity [71]-[72].

### 4.4. Limitations and Future Directions

This study investigates vocal stigma, for the very first time, in the context of social computing with expected technical challenges. To start, our vocal-stigma specific questionnaire was deployed to Canadians only for this initial round of analysis. Additional data from other countries can be collected to evaluate the sensitivity and specificity of VS-ABM or if additional organization or institutional factors (e.g., reimbursement system) will be needed to be included in future simulations. Second, our initial model results suggest the presence of a “*chaotic set*” in the population behavior. A more rigorous analysis of systems dynamics can be performed to evaluate when the system is in equilibrium or cycling in parameter space, which is out of the scope of this paper. Third, causal models can be considered to further examine the direction and causal relationships between components of IMB and stigma, since we primarily focused on correlations between these measures. Lastly, for the network analysis, we took a measured approach to establish a general trend for the population dynamics. However, more complex network measures such as information transmission and node similarity can be employed to characterize the details of stigma spread through the social network, instead of focusing on population averages in IMB and stigma.

## Conclusions

Stigma is a serious, intractable barrier that prevents vocal performers from seeking vocal healthcare. This study integrated empirical survey and network simulation approaches to quantify the relationship of social interaction and vocal stigma. VS-ABM computer models were built to numerically simulate peer interactions and track person-level changes in the IMB framework. Our results highlight that an individual’s social network plays an important role in the experience of vocal stigma. Especially for vocal performers, social interventions, e.g., close peer support, may help reduce high vocal stigma over time.

## Data Availability

The computational code and the research data that support the findings of this study are available from the corresponding author upon reasonable request.

## List of Abbreviations

ABM: Agent-based model
IMB: Information-Motivation-Behavioral Skills Model
LS/HS: Low stigma and High stigma groups
VP: Vocal performer

## Declarations

### Ethics approval and consent to participate

This study protocol (A09-B73-20A) was approved by the Institutional Review Board at McGill University. All participants of this study gave their informed, written consent.

### Consent for publication

Not applicable

### Contributions

**Aaron Glick:** Conceptualization (lead), Investigation (lead), Formal Analysis (equal), Writing – original draft (lead). **Colin Jones:** Formal Analysis (equal), Data Curation (lead), Writing – original draft (supporting). **Lisa Martignetti**: Investigation (supporting), Writing – review and editing (supporting). **Lisa Blanchette**: Investigation (supporting), Writing – review and editing (supporting). **Theresa Tova**: Investigation (supporting), Writing – review and editing (supporting). **Allen Henderson**: Investigation (supporting), Writing – review and editing (supporting). **Marc Pell:** Supervision (supporting), Conceptualization (supporting), Writing – review and editing (supporting). **Nicole Li-Jessen:** Supervision (lead), Conceptualization (supporting), Funding Acquisition (lead), Resources (lead), Writing – review and editing (lead).

## Acknowledgements

The authors would like to thank for all individuals who participated in the survey study.

## Supplementary Information

### Additional File 1: Study Questionnaire

The study questionnaire contained 64 items, grouped into 6 sections (sections A through F). Sections A, B, and C respectively pertained to demographics (6 items), occupation and training (3 items), and vocal health history (14 items). Section D measured predictors of help-seeking, and was divided into three subsections: DI, DM, and DB, respectively measuring participants’ levels of Information, Motivation, and Behavioral Skills. Section E measured experiences of vocal stigma. Finally, section F consisted of a single, open-ended item for feedback. Based on early piloting, we estimated the survey would take participants around 10-15 minutes to complete, which is a good length of survey to ensure low dropout rates[73]. The contents of each section are described below.

#### Survey Sections A and B: Demographics and Occupation

The questionnaire included a maximum of six items for demographics (including one item that was only presented to some participants, based on responses to previous items), and three items for occupational variables. Together, these nine items evaluated possible drivers and facilitators of stigma as well factors which could otherwise influence survey results (e.g., level of education).

#### Survey Section C: Vocal Health History

Vocal health variables were assessed by 14 items, including the complete Voice Handicap Index-10 (VHI-10). The VHI-10 is a well-validated [74] clinical tool commonly used to measure the impact that voice disorders have on quality of life. Two additional questions: one in which participants were asked when they had most recently experienced a voice disorder, and one which asked how frequently they experience voice disorders (with “never” being an available response for both). For participants who indicated that they have or previously had a voice disorder, two items ask whether they sought professional help, and if so, from which professionals.

#### Survey Section D: IMB Scales for Predicting Help-Seeking

All items in section D used a five-point, Likert scale format from 0 to 4, in line with the VHI-10. The scales were labeled as 0 = strongly disagree, 1 = disagree, 2 = neutral, 3 = agree, and 4 = strongly agree except Information. In all three sections, a participant’s likelihood of seeking help was represented as the sum of their scores on all ten items in the scale. This sum was a number between 0 and 40, where 40 represented the highest likelihood of seeking help and 0 the lowest. To reduce the impact of response bias, some items were reverse-coded, but item clarity was prioritized above creating a fully balanced scale [75].

##### Section D-I: Information Scale

Items in the Information scale were selected to assess participants’ knowledge and beliefs about the following five facets of voice and vocal health: (a) vocal health issues (items DI-2 and 10), (b) vocal health resources (DI-4 and 5), (c) voice care (DI-1 and 3), (d) vocal anatomy and physiology (DI-6 and 7), and (e) signs and symptoms of voice disorders (DI-8 and 9). Of these, DI-3, 4, 5, and 10 were adapted from Braun-Janzen and Zeine [76], DI-2 was adapted from Jung et al. [77], and the rest, DI-1, 6, 7, 8, and 9, were written for this study based on information in Colton et al. [78]. Response categories for Information items were labeled to indicate perceived truthfulness of the statements: 0 = definitely false, 1 = probably false, 2 = not sure, 3 = probably true, and 4 = definitely true. There were four reverse-coded items in the Information section.

##### Section D-M: Motivation Scale

Items in the Motivation scale were selected to assess participants’ attitudes and normative beliefs toward help-seeking for voice disorders. Items DM-1, 2, 3, 4, and 9 were either based on or taken directly from Gilman et al. [79]; item DM-10 was adapted from Christopher [80]; item DM-5 was adapted from Jung et al. [77]; and items DM-6, 7, and 8 were adapted from Fischer and Farina [81]. There were five reverse-coded items in this section.

##### Section D-B: Behavioral Skills Scale

Items in the Behavioral Skills scale were selected to assess participants’ intentions to seek help. Items DB-1 and 2 were adapted from the Mental Help-Seeking Intentions Scale, items DB-3 through 6 were adapted from the Intentions of Seeking Counselling Inventory, and DB-7 through 10 were adapted from the General Help-Seeking Questionnaire, all reviewed by Hammer and Spiker [82]. There was one reverse-coded Behavioral Skills item. This imbalance in reverse-coded items rises from a lack of reversed items in the source materials.

##### Survey Section E: Stigma Scale

Section E used the same Likert format as section D-M and D-B. The ten items making up the stigma scale were selected to assess participants’ perceptions of both self-stigma (items ES-1 through ES-5), and social-stigma (items EO-1 through EO-5). Items ES-1, 2, 3, and 5 were adapted from Vogel et al. [83], while ES-4 was written for this study based on Rosen et al. [84]. Items EO-1, 2, 3, and 5 were based on Clough et al. [85], and EO-4 was based on discussions of VPs’ reputation found in, among others, Bradshaw and Cooper [86], Sataloff et al. [87], and Sloggy et al. [88]. The stigma scale included four reverse-coded items. As with each of the I, M, and B sections, the level of stigma experienced by a participant was measured as the sum of the ten scores.

#### Survey Section F: Open-Ended Feedback

After answering all the other items, participants could share perspectives on voice disorders and vocal stigma that they felt were not addressed by the rest of the questionnaire.

*<u>Special Notes</u>: Below is a reproduction of the content in the final survey. Text in italics is for the reader’s information only and was not visible to participants. Questions marked with a * were reverse-coded*.

### WELCOME MESSAGE

#### Vocal Illness Experiences

Welcome to this survey about people’s experiences with vocal illnesses. We’ll start with a few simple questions to make sure you fit our study’s criteria. After that, there will be a consent form, followed by a survey.

You can save your progress and return at any time, but you can’t go back to previous questions after you have moved to the next page.

Click next to continue.

### PARTICIPANT SCREENING

*X1* **Do you have a Canadian Address?**

a. Yes
b. No
*X2* **Are you between the ages of 20 and 65?**

a. Yes
b. No
*X3* **Has your voice ever been affected by one of the following conditions:**

- **Cancer in the head/neck**
- **Stroke**
- **Parkinson’s Disease, ALS, or other neurodegenerative conditions**
- **Physical trauma to the throat, neck, or head (e.g., caused by car accident)**

a. Yes
b. No

*Participants who answered X1, yes; X2, yes; X3, no were able to continue to the consent section. Otherwise, they saw the following message:*

“Thank you for responding to our survey. Unfortunately, you don’t quite fit for what we’re trying to study. Feel free to share our study if you know anyone who does, so they can enter the draw for a gift card!”

### CONSENT SECTION

*Consent split* Are you a professional singer or actor (i.e., you make at least some income via performance)?

a. Yes
b. No

*Participants who answered “yes” became part of the performer group, while participants who answer “no” became part of the control group. Their respective consent forms were presented here*.

*Participants in both groups who did consent to participate were able to proceed to the rest of the study.*

A. – Demographics

- *A-1* **What gender do you identify as?**

a. Male
b. Female
c. I prefer not to answer
d. Other: ____________
- *A-2* **Are you a member of a visible minority?**

a. No
b. Yes
c. I prefer not to answer
- *A-3* **What is your age? (Years) ____________**
- *A-4* **What are the first three characters of your Canadian post-code? ____________**
- *A-5* **What is the highest level of education you have completed?**

a. High school diploma
b. Apprenticeship or trades certificate/diploma
c. College or CEGEP degree/diploma, or university degree lower than bachelor’s
d. Bachelor’s degree
e. Graduate degree (master’s or doctorate)
f. None of the above

*Participants who responded to A-5 with any answer other than (a) or (f) were shown item A-5.1:*

*A-5.1* **In which area(s) did you obtain a degree/diploma/certificate?**

Check all that apply

a. Performance (e.g., music, acting, dance…)
b. Healthcare
c. Biology or Physiology
d. Other
*B - Occupation*

*B-1* Which of these statements about your occupation best describe you?

a. I am a professional actor (including theatre, film/TV, Voice Actor; not including musical theatre/opera).
b. I am a professional singer (including concerts, recordings, staged productions; solo and ensemble).
c. I am not a professional vocal performer.
*B-2* How much voice training have you received? (Training includes any lessons, classes, or coaching with a focus on learning voice skills for spoken or sung performance.)

a. None
b. Less than 1 year
c. 3 years or less
d. 5 years or less
e. More than 5 years
*B-3* Approximately how much of your income is made by working as a singer or actor in an average year (pre-pandemic)?

a. None or almost none
b. Less than half
c. About half
d. More than half
e. All or almost all
f. I prefer not to answer
C - Vocal Health

**Please read the following information about voice disorders before continuing.**

**Voice disorders are a wide range of conditions that impact a person’s voice in various ways, including the tone, pitch, loudness, and more. A voice disorder is not the same a speech disorder (which impacts your ability to speak fluently and accurately, such as a stutter or a lisp).**

**For this study, <u>a voice disorder</u> refers to any disturbance to how your voice normally functions or sounds, in a way that interferes with your daily conversation and/or your professional work as a performer.**

Exceptions: (the following would <u>not</u> be considered a voice disorder):

- **The problem resolves on its own within 1-2 days and does not come back regularly.**
- **The problem is related to a brief illness such as a cold or flu, and voice symptoms resolve at a similar time to other symptoms**.
C-1 When was the last time you had a voice disorder?

a. I have never had a voice disorder
b. I currently have a voice disorder
c. Within the past month
d. Within the past year
e. More than a year ago

*Participants who responded to C-1 with any answer other than (a) were shown item C-1.1:*

*C-1.1* **Did you approach a professional to help identify, overcome, or cope with this issue?** (In this question, a professional can include anyone whose job includes helping people with their voice or other health issues. Possible examples include: doctor, speech-language pathologist, voice teacher/coach, massage therapist, psychologist, acupuncturist, etc.)

a. No
b. Yes

*Participants who responded to C-1.1 with (b), were shown item C-1.1.1:*

*C-1.1.1* What type of professional did you seek help from? (You may list several)

____________
____________
____________
____________
____________
C-2 **How often have you experienced a voice disorder?**

a. Never
b. Once every few years
c. Once or twice every year
d. Once or twice every three months
e. Once or twice every month
f. Almost all the time
C-3 **These are statements that many people have used to describe their voices and effects of their voices on their lives. Choose the response that indicates how frequently you have the same experience.** 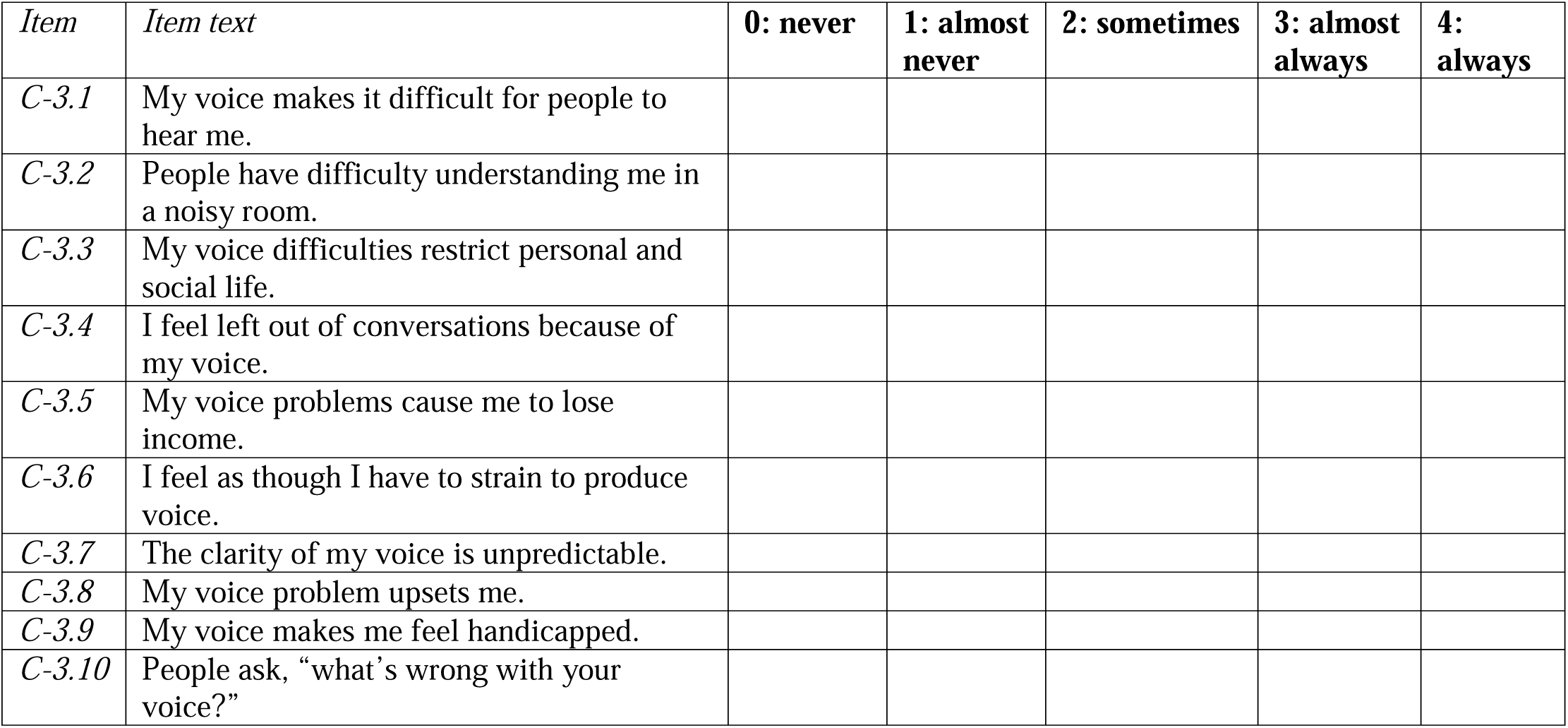

***D – Likelihood of Seeking Help***

***DI: Information***

**Please rate the following statements according to how confident you are that they are true: 0 = definitely false; 1 = probably false; 2 = not sure; 3 = probably true; 4 = definitely true**

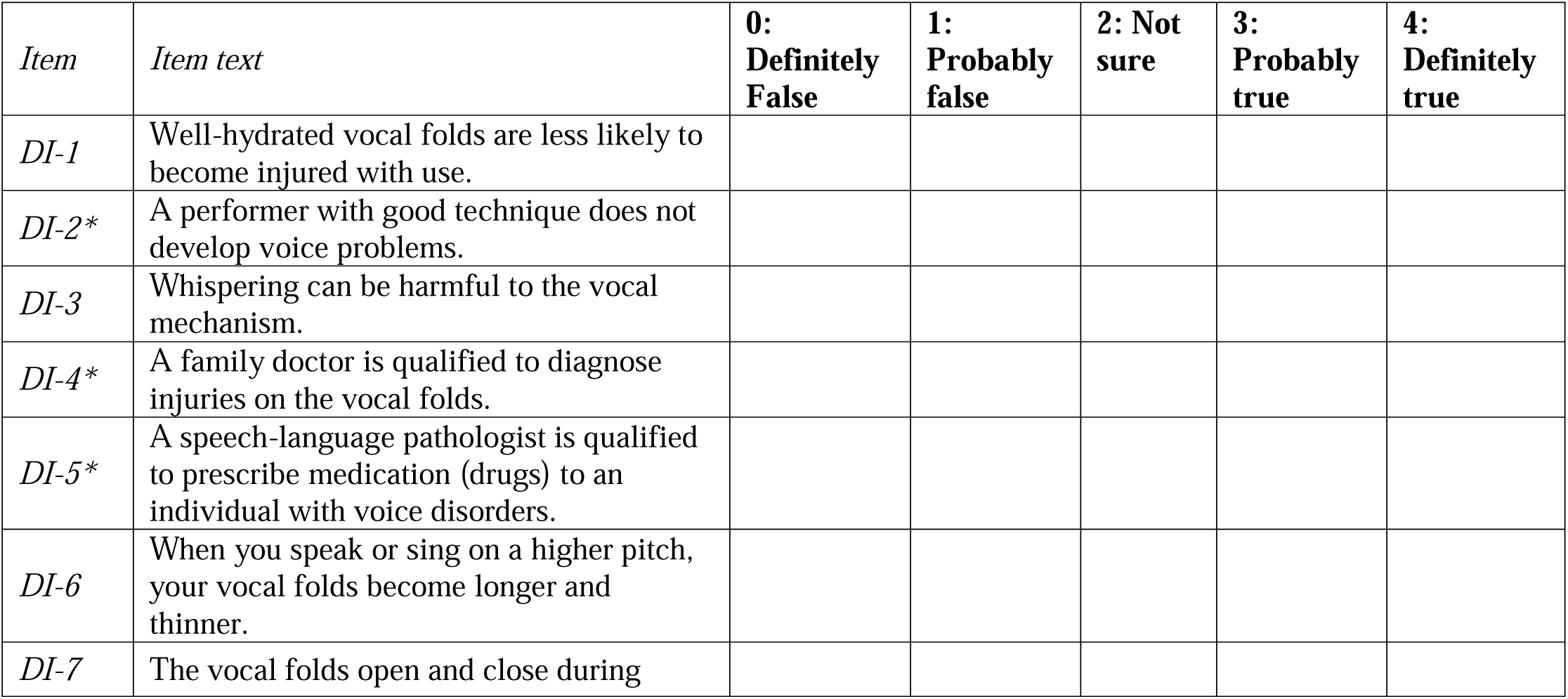

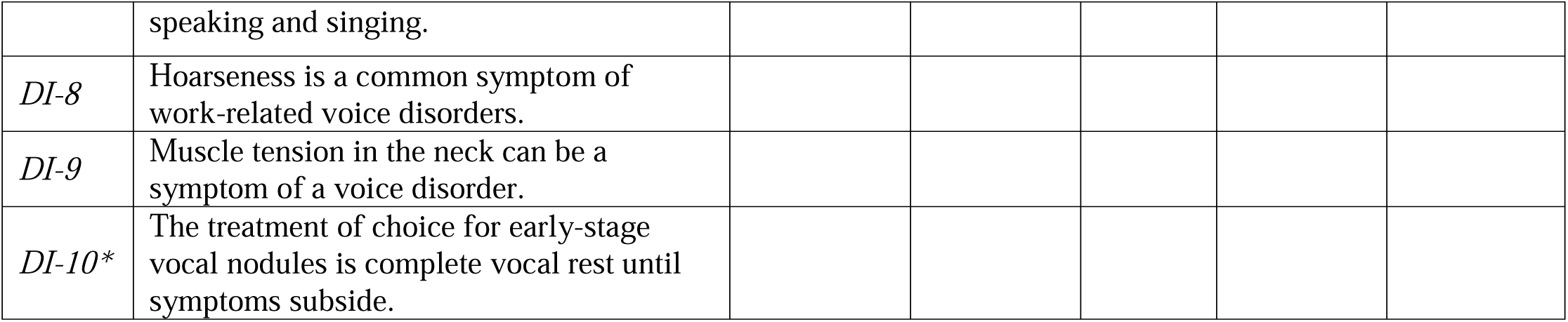

***DM: Motivation***

**Please rate the statements below on a scale of 0 – 4, where:**

**0 = strongly disagree; 1 = disagree; 2 = neutral; 3 = agree; 4 = strongly agree**

| *Item* | *Item text* | **0: strongly disagree** | **1: disagree** | **2: neutral** | **3: agree** | **4: strongly agree** |
| --- | --- | --- | --- | --- | --- | --- |
| *DM-1\** | I do not see a specialist when I have voice problems because I am afraid of what may be found. |  |  |  |  |  |
| *DM-2\** | I may not seek health care for my voice problem because I anticipate or experience difficulty with accessing public health services. |  |  |  |  |  |
| *DM-3\** | I have anxiety/fear about going to a specialist for problems with my voice. |  |  |  |  |  |
| *DM-4* | My voice is an important part of my profession. |  |  |  |  |  |
| *DM-5* | Speech therapy is a helpful treatment for voice problems. |  |  |  |  |  |
| *DM-6* | A person with a voice disorder is not likely to solve it alone. |  |  |  |  |  |
| *DM-7\** | A person should work out their own problems; getting voice therapy would be a last resort. |  |  |  |  |  |
| *DM-8\** | There is something admirable in the attitude of someone who is willing to cope with their voice difficulties without resorting to professional help. |  |  |  |  |  |
| *DM-9\** | I may not seek health care for my voice problem because I anticipate or experience difficulty with affording private health services. |  |  |  |  |  |
| *DM-10* | If I were experiencing a voice problem, my work colleagues would think I should seek professional vocal health services. |  |  |  |  |  |

**DB: Behavioral Skills**

**Please rate the statements below on a scale of 0 – 4, where:**

**0 = strongly disagree; 1 = disagree; 2 = neutral; 3 = agree; 4 = strongly agree**

| *Item* | *Item text* | **0: strongly disagree** | **1: disagree** | **2: neutral** | **3: agree** | **4: strongly agree** |
| --- | --- | --- | --- | --- | --- | --- |
| *DB-1* | If I had a vocal health concern, I would intend to seek help from a vocal health professional. |  |  |  |  |  |
| *DB-2\** | If I was experiencing a voice disorder, I would try to solve it on my own rather than seeking help. |  |  |  |  |  |

**The next 4 items fill in the blank of the following sentence:**

**“If I was experiencing a voice disorder, I would be likely to seek help from____________.”**

**The next 4 items fill in the blank of the following sentence:** **“If I was experiencing a voice disorder, I would be likely to seek help from \_\_\_\_\_.”**
| *Item* | *Item text* | **0: strongly disagree** | **1: disagree** | **2: neutral** | **3: agree** | **4: strongly agree** |
| --- | --- | --- | --- | --- | --- | --- |
| *DB-3* | ...a family doctor/general physician. |  |  |  |  |  |
| *DB-4* | ...a laryngologist/ENT (Ear, Nose, and Throat doctor) |  |  |  |  |  |
| *DB-5* | ...a speech-language pathologist. |  |  |  |  |  |
| *DB-6\** | ...an alternative medicine practitioner (e.g., acupuncture, reiki, homeopathy) |  |  |  |  |  |

**For the purposes of the rest of the questionnaire, the term “vocal health professional” will refer to laryngologists/ENT (Ear, Nose, and Throat doctors), and speech-language pathologists.**

**The next 4 items fill in the blank of the following sentence:**

“I would be likely to seek help from a vocal health professional if ____________.”

| *Item* | *Item text* | **0: strongly disagree** | **1: disagree** | **2: neutral** | **3: agree** | **4: strongly agree** |
| --- | --- | --- | --- | --- | --- | --- |
| *DB-7* | ... my voice sounded hoarse or rough. |  |  |  |  |  |
| *DB-8* | ... my voice often felt tired after use. |  |  |  |  |  |
| *DB-9* | ... using my voice felt painful or uncomfortable. |  |  |  |  |  |
| *DB-10* | ... I had to cancel a performance/contract because of voice problems. |  |  |  |  |  |

***E - Experiences of Stigma***

**Please rate the statements below on a scale of 0 - 4, where:**

**0 = strongly disagree; 1 = disagree; 2 = neutral; 3 = agree; 4 = strongly agree**

*Items numbered as EO pertain to social-stigma; items numbered as ES pertain to self-stigma*

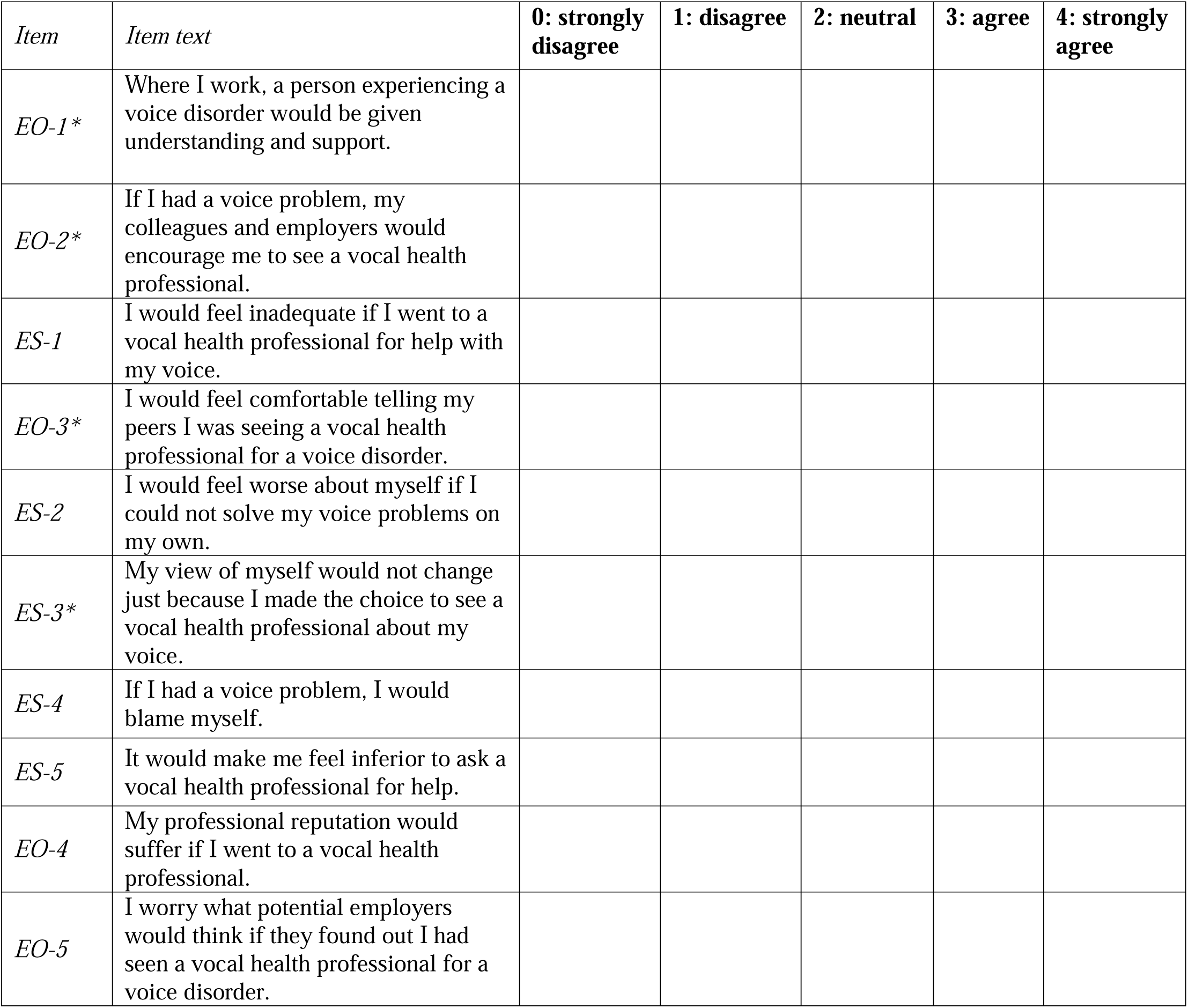

**Reminder: for this survey, “vocal health professional” refers to laryngologists/ENTs (Ear, Nose, and Throat doctors), and speech-language pathologists.**

***F – Feedback***

*F-1* If there is anything you would like to say about stigma and voice disorders that was not covered by this questionnaire, you can tell us here:

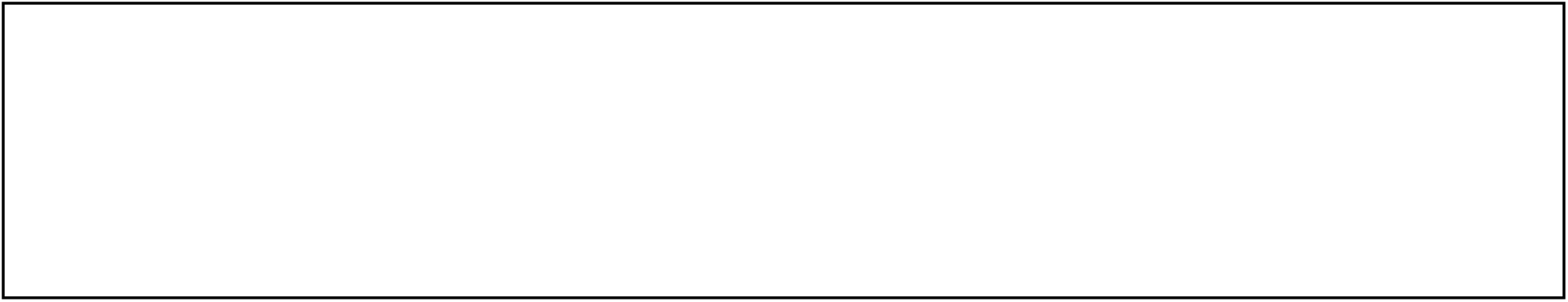

## Additional File 2: Study participant demographics

**Supplementary Table 2.1.** **Participant Demographics**. Data are presented as response frequencies except the age variable as group means.

| Variable | Category | Vocal Performers |  | Controls |  | *p* |
| --- | --- | --- | --- | --- | --- | --- |
| **Gender identity** |  |  |  |  |  | .835<sup>a</sup> |
|  |  | *n* | % | *n* | % |  |
|  | Male | 64 | 32 | 65 | 32 |  |
|  | Female | 130 | 65 | 132 | 65 |  |
|  | Other | 4 | 2 | 6 | 3 |  |
|  | Prefer not to answer | 2 | 1 | 0 | 0 |  |
| **Visible minority status** |  |  |  |  |  | .001<sup>a</sup> |
|  |  | *n* | % | *n* | % |  |
|  | Not a visible minority | 156 | 78 | 135 | 66.5 |  |
|  | Visible minority | 37 | 18.5 | 68 | 33.5 |  |
|  | Prefer not to answer | 7 | 3.5 | 0 | 0 |  |
| **Rural vs non-rural** |  |  |  |  |  | .973<sup>a</sup> |
|  |  | *n* | % | *n* | % |  |
|  | Rural | 10 | 5 | 10 | 4.9 |  |
|  | Non-Rural | 190 | 95 | 193 | 94.6 |  |
| **Education level** |  |  |  |  |  | .288<sup>a</sup> |
|  |  | *n* | % | *n* | % |  |
|  | High school diploma | 18 | 9 | 26 | 12.8 |  |
|  | Apprenticeship or trades certificate/diploma | 10 | 5 | 4 | 2 |  |
|  | College or CEGEP degree/diploma, or university degree lower than bachelor's | 33 | 16.5 | 29 | 14.3 |  |
|  | Bachelor's degree | 92 | 46 | 102 | 50.2 |  |
|  | Graduate degree (master's or doctorate) | 47 | 23.5 | 41 | 20.2 |  |
|  | None of the above | 0 | 0 | 1 | 0.5 |  |
| **Degree area** |  |  |  |  |  | <.001<sup>a</sup> |
|  |  | *n* | % | *n* | % |  |
|  | Performance | 124 | 62 | 2 | 1 |  |
|  | Health sciences | 10 | 5 | 37 | 18.2 |  |
|  | Both performance and health sciences | 6 | 3 | 0 | 0 |  |
|  | Other area(s) only | 42 | 21 | 137 | 67.5 |  |
|  | N/A<sup>c</sup> | 18 | 9 | 27 | 13.3 |  |
| **Age** |  |  |  |  |  | <.001<sup>b</sup> |
|  |  | **Years** |  | **Years** |  |  |
|  | Mean | 43.68 |  | 33.76 |  |  |
|  | SD | 12.56 |  | 10.29 |  |  |

<sup>a</sup>Results of a chi-squared test of independence between participant groups.
<sup>b</sup>Results of a Student's t-test between participant groups.
<sup>c</sup>N/A= participants to whom this item was not presented because they reported their education level as "high-school diploma" or "none of the above."

**Supplementary Table 2.2.**
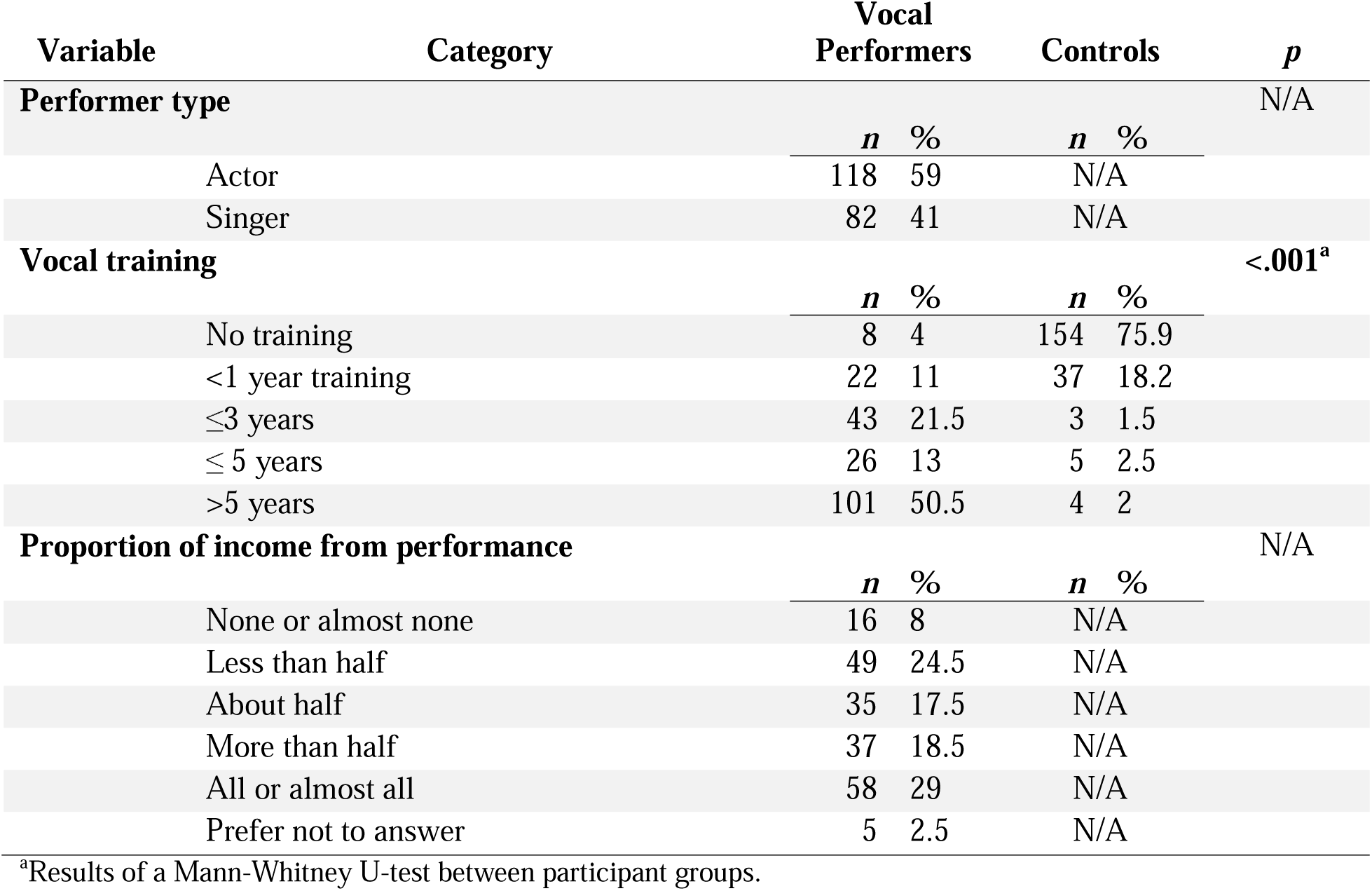
**Occupation and Training**. Data are presented as response frequencies.

**Supplementary Table 2.3.**
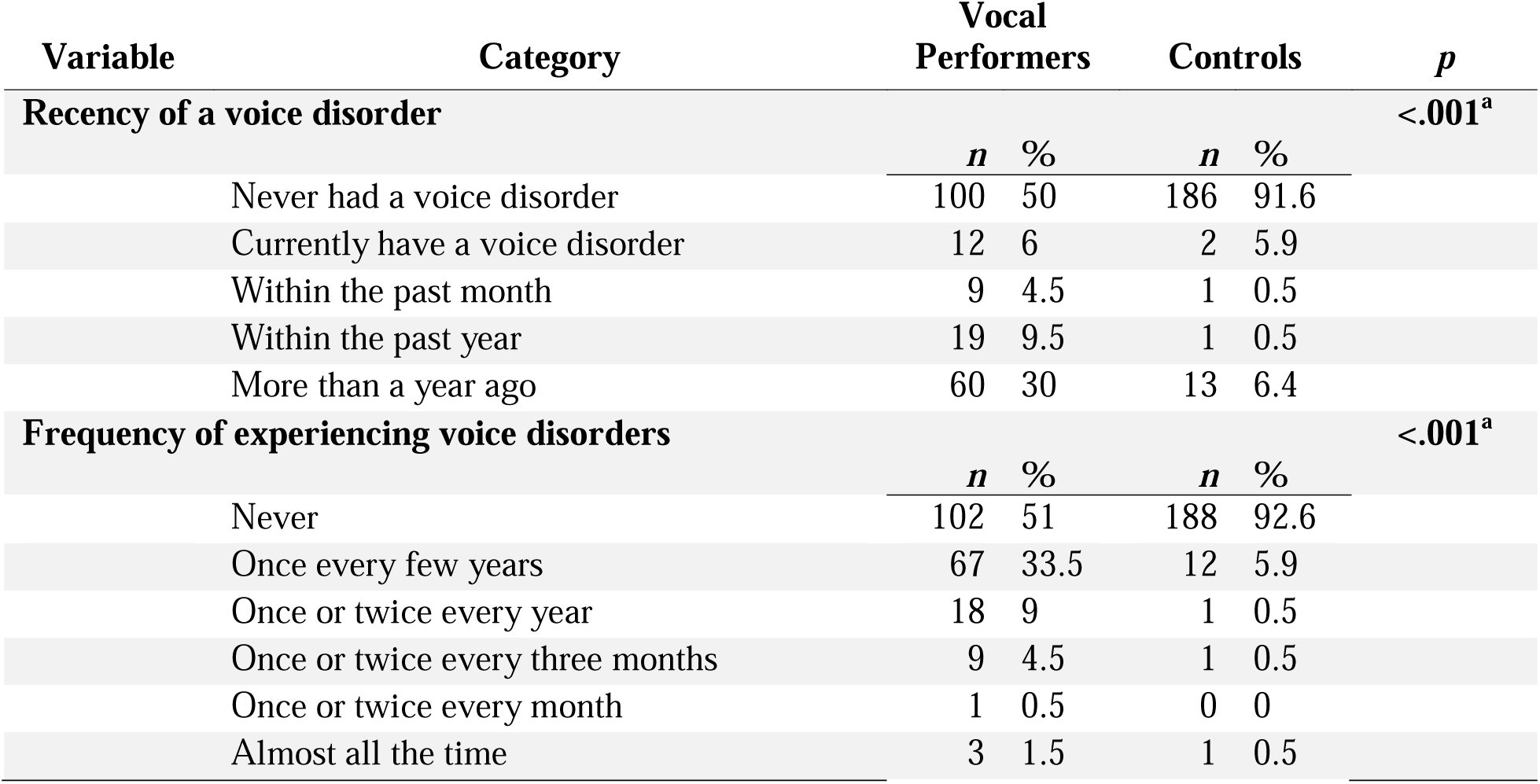
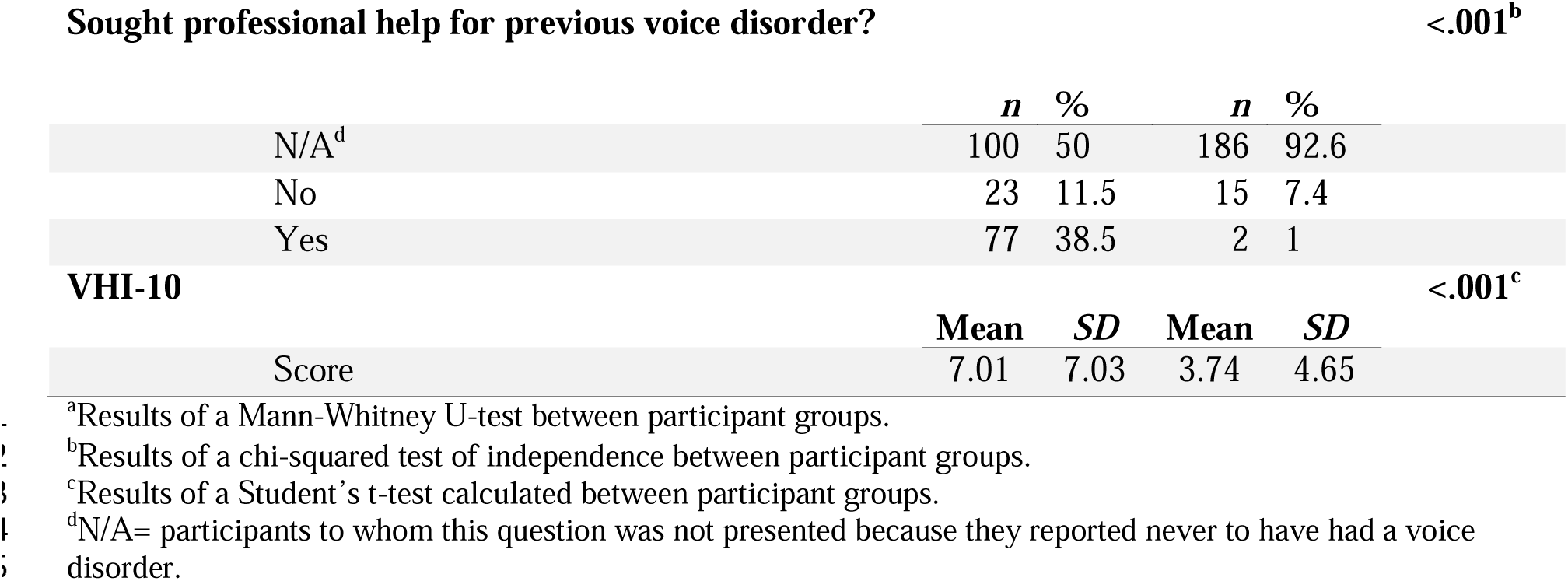
**Vocal Health Profiles.** Data are presented as response frequencies except VHI-10 score in group means.

## Additional File 3: Questionnaire results for VP and control group

**Table 3.1.** **Means and standard deviations (SD) of stigma experience scores between vocal performers and controls. * < 0.05**

| Scores | Vocal Performers | Controls | t(401) | p |
| --- | --- | --- | --- | --- |
| **Total stigma (max: 40)** | 11.15 (6.68) | 9.74 (5.84) | 2.25 | .025* |
| **Self-stigma (max: 20)** | 5.20 (3.76) | 4.88 (3.50) | 0.88 | .379 |
| **Social-stigma (max: 20)** | 5.95 (3.85) | 4.86 (2.96) | 3.18 | .002* |

**Table 3.2.** **Means and standard deviations (SD) of IMB scores between vocal performers and controls. * < 0.05**

| Scores | Vocal Performers | Controls | t(401) | p |
| --- | --- | --- | --- | --- |
| **Information (max: 40)** | 26.88 (4.64) | 23.48 (2.87) | 8.87 | <.001* |
| **Motivation (max: 40)** | 30.60 (5.00) | 28.01 (4.99) | 5.19 | <.001* |
| **Behavioral skills (max: 40)** | 28.69 (4.79) | 27.52 (4.96) | 2.41 | .017* |

**Table 3.3.** **Correlations for help-seeking and stigma relationship from the vocal performer group**

|  | Information | Motivation | Behavioral Skills | Total Stigma | Self-stigma | Social-stigma |
| --- | --- | --- | --- | --- | --- | --- |
| **Information** | 1.00 | - | - | - | - | - |
| **Motivation** | .247*** | 1.00 | - | - | - | - |
| **Behavioral skills** | .225*** | .534*** | 1.00 | - | - | - |
| **Total stigma** | -.09 | -.49*** | -.28 | 1.00 | - | - |
| **Self-stigma** | -.079 | -.441*** | -.231*** | - | 1.00 | - |
| **Social-stigma** | -.079 | -.410*** | -.267*** | - | .540*** | 1.00 |

Significance levels reported as \*\* $p = .01$ \*\*\* $p < .001$

**Table 3.4.** **Correlations for help-seeking and stigma relationship from the control group**

|  | Information | Motivation | Behavioral Skills | Total Stigma | Self-stigma | Social-stigma |
| --- | --- | --- | --- | --- | --- | --- |
| **Information** | 1.00 | - | - | - | - | - |
| **Motivation** | .191** | 1.00 | - | - | - | - |
| **Behavioral skills** | .050 | .534*** | 1.00 | - | - | - |
| **Total stigma** | -.09 | -.59*** | -.46*** | 1.00 | - | - |
| **Self-stigma** | -.078 | -.534*** | -.391*** | - | 1.00 | - |
| **Social-stigma** | -.086 | -.542*** | -.450*** | - | .645*** | 1.00 |

Significance levels reported as \*\* $p = .01$ \*\*\* $p < .001$

**Table 3.5.** **Linear regression predictions for help-seeking and stigma relationship from the performer group**

|  | Behavioral skills | Self-stigma | Social-stigma |
| --- | --- | --- | --- |
| **Information** | 0.232 | 0.064 [6.911] | 0.066 [7.720] |
| **Motivation** | 0.511 | 0.267 [15.341] | 0.316 [15.604] |
| **Behavioral skills** | - | 0.181 [10.400] | 0.215 [12.106] |
| **Self-stigma** | - | - | 1.805 [5.532] |

Regression terms for $y = mx + b$ reported as $m[b]$ with $x$ listed in rows and $y$ listed in columns

**Table 3.6.** **Means and standard deviations (SD) of IMB and Stigma values for demographic subgroups related to history of voice disorder and tendency to seek professional help. Bolded numbers are used to set up the five agent populations in the VS-ABM.**

|  |  | Information | Motivation | Behavioral Skills | Self-stigma | Social-stigma |
| --- | --- | --- | --- | --- | --- | --- |
| **1.** | General population (403) | **25.17 (4.20)** | **29.30 (5.15)** | **28.10 (4.90)** | **5.04 (3.63)** | **5.40 (3.47)** |
| **2.** | All Controls (203) | **23.48 (2.87)** | **28.01 (4.99)** | **27.52 (4.96)** | **4.88 (3.50)** | **4.86 (2.96)** |
| **2.a** | Controls no history of voice disorder (186) | 23.49 (2.90) | 28.35 (4.92) | 27.79 (4.75) | 4.78 (3.54) | 4.73 (2.99) |
| **2.b** | Controls with history of voice disorder (17) | 23.29 (2.59) | 24.29 (4.18) | 24.59 (6.31) | 5.94 (2.94) | 6.23 (2.25) |
| **2.b.i** | Controls with history who did not seek help (15) | 22.8 (2.31) | 23.67 (3.89) | 23.93 (6.44) | 6.2 (3.05) | 6.53 (2.20) |
| **2.b.ii** | Controls with history who sought help (2) | 27.00 (1.41) | 29.00 (4.24) | 29.50 (0.71) | 4.00 (0) | 4.00 (1.41) |
| **3.** | All VPs (200) | **26.88 (4.63)** | **30.60 (5.00)** | **28.69 (4.79)** | **5.2 (3.76)** | **5.95 (3.85)** |
| **3.a** | VPs no history of voice disorder (100) | 26.37 (3.95) | 31.41 (4.41) | 28.93 (4.41) | 4.29 (3.41) | 5.25 (3.29) |
| **3.b** | VPs with history of voice disorder (100) | 27.39 (5.20) | 29.78 (5.43) | 28.45 (5.15) | 6.11 (3.88) | 6.64 (4.24) |
| **3.b.i** | VPs with history who did not seek help (23) | **28.96 (4.41)** | **27.17 (5.17)** | **25.70 (4.88)** | **6.52 (4.13)** | **7.39 (4.35)** |
| **3.b.ii** | VPs with history who sought help (77) | **26.92 (5.35)** | **30.34 (5.41)** | **29.27 (4.97)** | **5.99 (3.82)** | **6.42 (4.21)** |

## Additional File 4: Agent-based model rule for Stigma and IMB model

**Supplementary Table 4.1.**
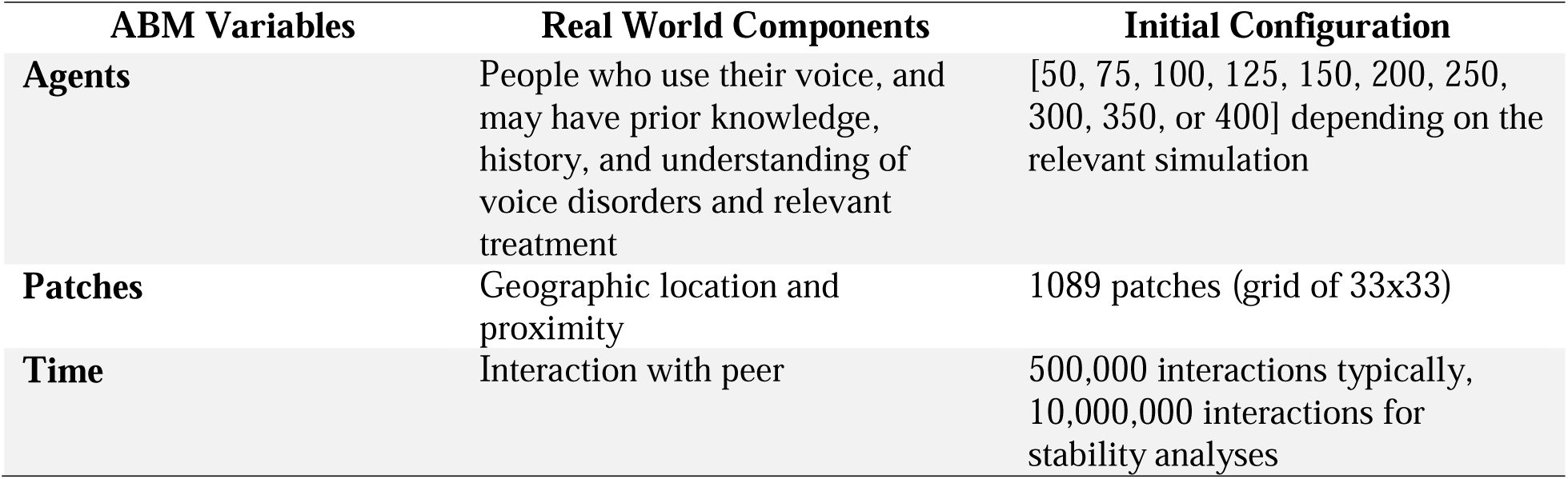
**Initial Configuration of VS-ABM**.

**Supplementary Table 4.2.**
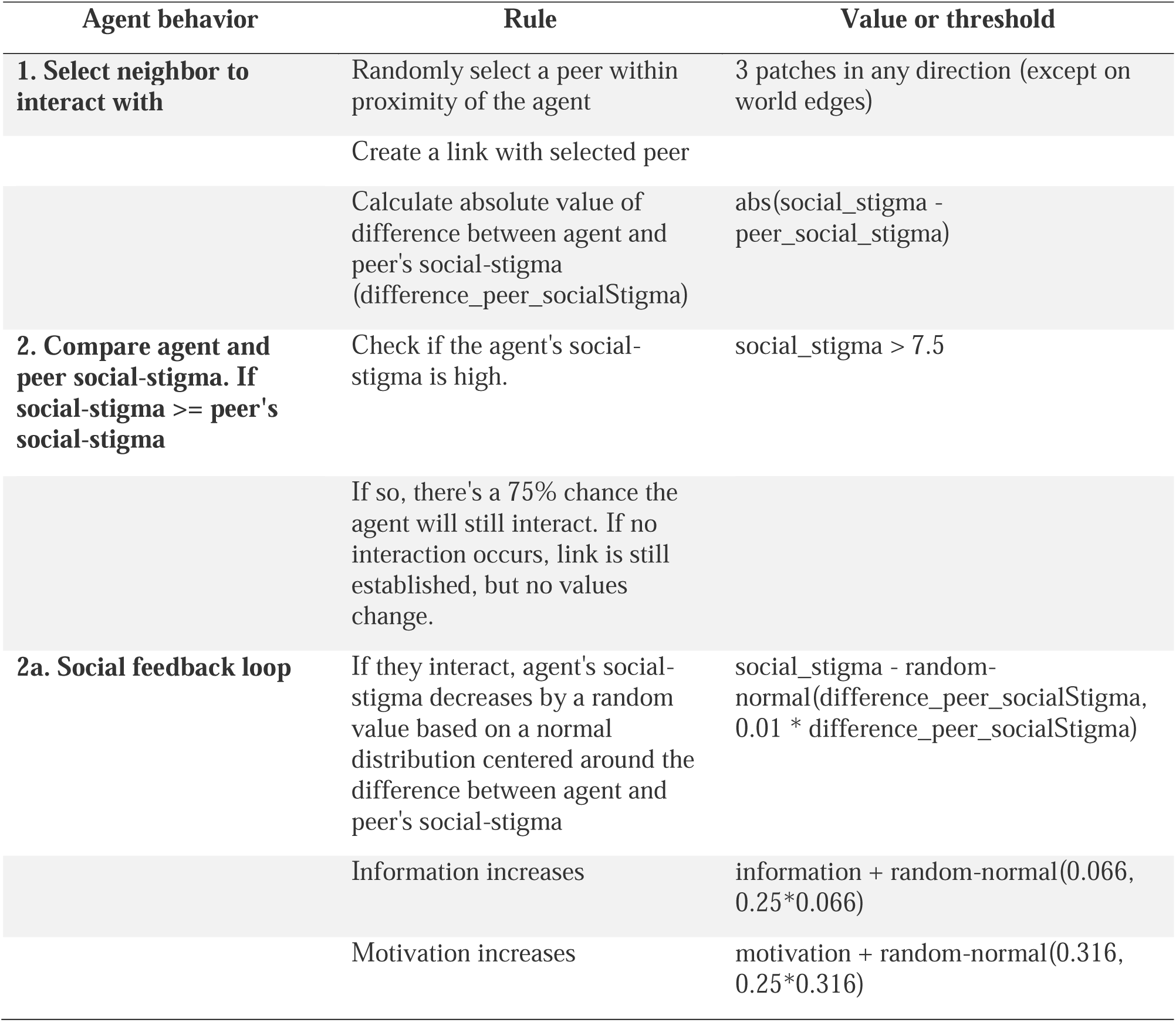
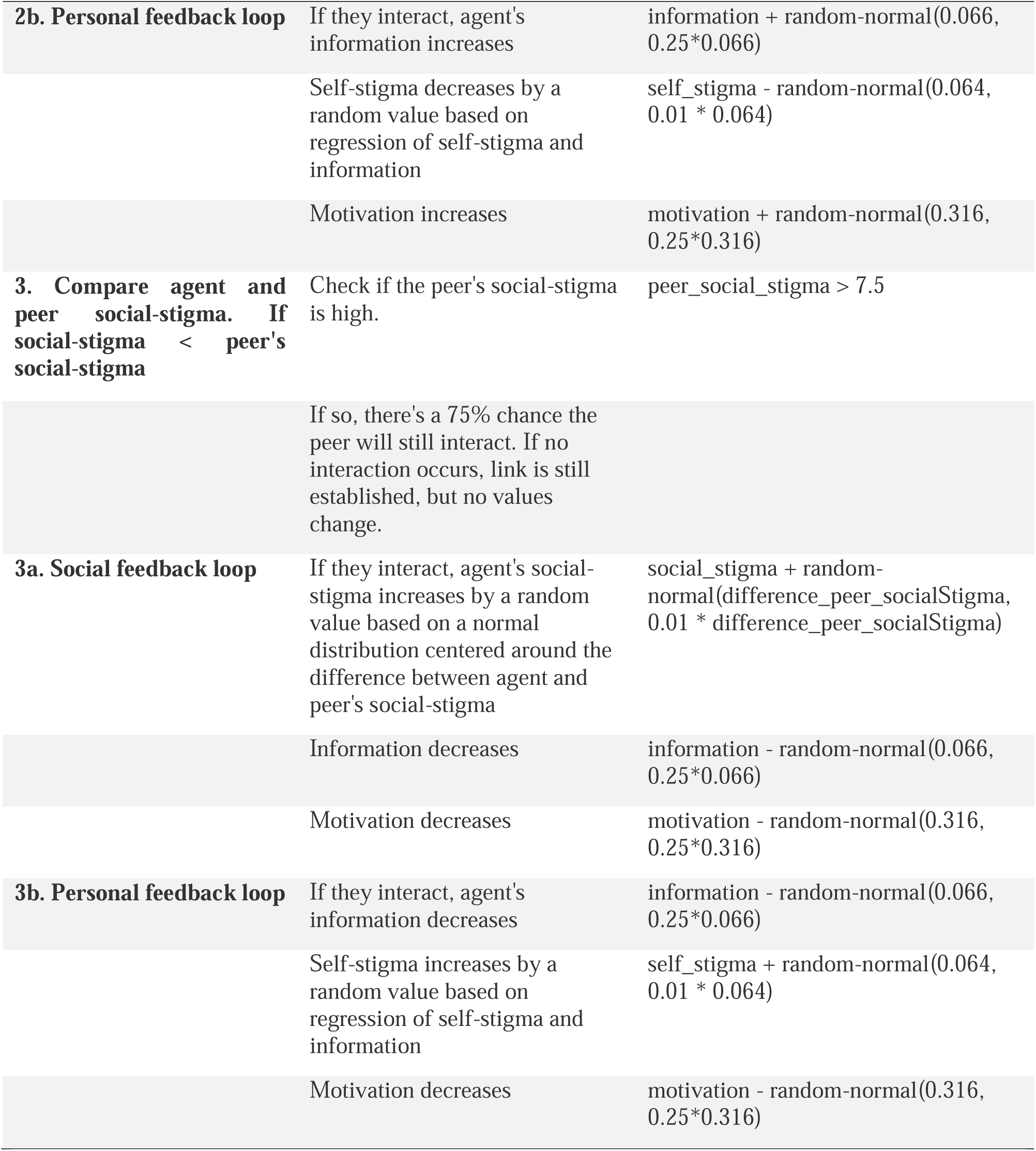
**Rules of VS-ABM**

**Supplementary Table 4.3.** **Initial Values for Analysis Groups of VS-ABM Simulation Experiments**

| Group | Real World Components | Mean | Standard Deviation |
| --- | --- | --- | --- |
| **Constant Parameters (based on VP population)** |  |  |  |
|  | Age | 43.68 | 12.56 |
|  | Information | 26.88 | 4.64 |
|  | Motivation | 30.60 | 5.00 |
|  | Behavior Skills | 28.69 | 4.79 |
|  | Self-Stigma | 5.20 | 3.76 |
| **Variable Parameters (100 simulations run for each set of variable parameters 1 and 2)** |  |  |  |
| **1a. Network size: 50** | Number of agents | 50 |  |
| **1b. Network size: 100** | Number of agents | 100 |  |
| **1c. Network size: 150** | Number of agents | 150 |  |
| **1d. Network size: 200** | Number of agents | 200 |  |
| **1e. Network size: 250** | Number of agents | 250 |  |
| **1f. Network size: 300** | Number of agents | 300 |  |
| **1g. Network size: 350** | Number of agents | 350 |  |
| **1h. Network size: 350** | Number of agents | 400 |  |
| **2a. non-VP Group** | Social-Stigma | 4.86 | 3.756 |
| **2b. General Population** | Social-Stigma | 5.46 | 3.756 |
| **2c. VP Group** | Social-Stigma | 5.95 | 3.756 |
| **2d. VP Group, high risk, with stigma mitigation** | Social-Stigma | 6.42 | 3.756 |
| **2e. VP Group, high risk, with no stigma mitigation** | Social-Stigma | 7.39 | 3.756 |

